# Diagnostic serial interval as an alternative measure of clinical serial interval using ancestral COVID-19 waves in Hong Kong and mainland China

**DOI:** 10.1101/2025.10.15.25338050

**Authors:** M. Pear Hossain, Dongxuan Chen, Amy Yeung, Dillon C. Adam, Wey Wen Lim, Yiu Chung Lau, Eric H. Y. Lau, Jessica Y. Wong, Faith Ho, Huizhi Gao, Lin Wang, Zhanwei Du, Peng Wu, Benjamin J. Cowling, Sheikh Taslim Ali

## Abstract

To estimate the reproductive numbers *(R*_*t*_), it is essential to infer generation time, which is often approximated by serial interval *(SI)*. The *SI* based on clinical outcomes is not free from recall biases and even such clinical information is not always available. We defined diagnostic serial interval *(SI*_*d*_), as a potential alternative metric and compared with traditional *SI*. We analyzed confirmed COVID-19 cases from three ancestral waves in Hong Kong and the first wave in mainland China. Using Bayesian methods, we inferred the distributions of effective *SI* and *SI*_*d*_, along with onset-to-reporting delays, and compared the resulting estimates. The distributions of *SI* and *SI*_*d*_ were comparable across waves, with shorter means observed in *SI*_*d*_. Reporting delays for infectors (*d*_1_) were longer than these for infectees (d_2_), which was identified as a key factor influencing the temporal variation in *SI*_*d*_. The PHSMs, case profile and demography were also found to be significant factors of *SI*_*d*_. Time-varying *R*_*t*_ estimates derived from both *SI* and *SI*_*d*_ were comparable, with median absolute differences ranging from 0.12 to 0.19. Therefore, *SI*_*d*_ shows potentials as an alternative metric to *SI* for estimating *R*_*t*_ in assessing COVID-19 transmission dynamics can be extended for other respiratory viruses.

## Introduction

Along with several epidemiological parameters, the inference of generation time (*GT*, the interval between successive infections in the transmission chain) and clinical serial interval (*SI*, the interval between successive onsets in the transmission chain) distributions is crucial for understanding the transmission dynamics of directly transmitted infectious diseases, including COVID-19^1–6^. The *GT* distribution is especially essential for inferring effective or instantaneous reproduction numbers (*R*_*t*_, a measure of transmissibility defined as the average number of secondary cases produced by a primary case at time *t*), which is useful for informing real-time public health decision-making^7–11^. In practice, times of infection of individuals are rarely observed, making it challenging to infer the *GT*. Therefore, clinical process outcomes such as the *SI* distribution^12–14^ are often used as a proxy for the *GT* distribution. This can subsequently be used to estimate reproduction numbers *(R*_0_: basic and *R*_*t*_)^15–18^ and other measures of transmissibility, such as secondary attack rate, force of infection, and doubling time etc^19–22^.

However, it is not always possible to routinely track the data on clinical onsets for the cases, often subject to recall biases, especially during high community activity^23^. On the other hand, reporting times are frequently recorded with greater precision and are available for a larger proportion of cases. Further, the presence of pre-symptomatic transmissions may return negative *SI*, which is not realistic to approximate *GT*, a positive measure ^14^ Therefore, it is preferable to avoid these limitations when estimating *GT* for respiratory diseases based on diagnostic information rather than clinical outcomes. This provides us an opportunity to use *diagnostic serial interval* (*SI*_*d*_, the time between diagnostic reporting between cases in transmission pairs) as a comparable alternative measure for the clinical *SI* and hence a reliable proxy for *GT* under a stable surveillance system. *SI*_*d*_ relies only on reporting times, which may overcome many limitations of clinical *SI*, including recall bias^27^ (as tests are performed and reported by trained professionals). Additionally, *SI*_*d*_ can be estimated even for asymptomatic infections, which have been reported with varying proportions from 1% to over 50% for COVID-19^28^. The only previous study we are aware of reporting for COVID-19 in South Korea by retrospectively estimating *SI*_*d*_ as a measure of contact tracing effectiveness using very initial data in 2020^29^. No further studies have explicitly explored temporal characteristics of *SI*_*d*_ and associated potential factors to illustrate the utility and inter-parametric dependency of *SI* and *SI*_*d*_ when estimating time-varying *R*_*t*_. This study aimed to assess the similarity of *SI*_*d*_ with clinical *SI* (a proxy of the generation time), using line-list data for ancestral COVID-19 waves in Hong Kong and mainland China during 2020-2021.

## Results

We analyzed all locally diagnosed COVID-19 cases in Hong Kong reported during the first four waves until 23 March 2021. This included data from the second wave (1 March to 10 April 2020), the third wave (25 June to 8 September 2020), and the fourth wave (1 November 2020 to 23 March 2021), while excluding the first wave due to its predominantly imported cases. Additionally, similar line-list data was retrieved for mainland China, covering cases reported from 1 January to 29 February 2020 (Material and Method section). During the study period in Hong Kong, we were able to construct 2,433 transmission pairs, including 823 confirmed transmission pairs (47 in the second wave, 357 in the third wave, 355 in the fourth wave, and 64 pairs identified during inter-wave periods), with the remaining being probable/likely pairs. In mainland China, a total of 629 confirmed pairs were reconstructed in the first wave. The onset and report-based epi-curves were generated independently using the respective onset and reporting of case data, which showed a similar trend with a delay of approximately 5-10 days in the observed peak timings across the waves (Figure 1A-D).

**Figure 1.**
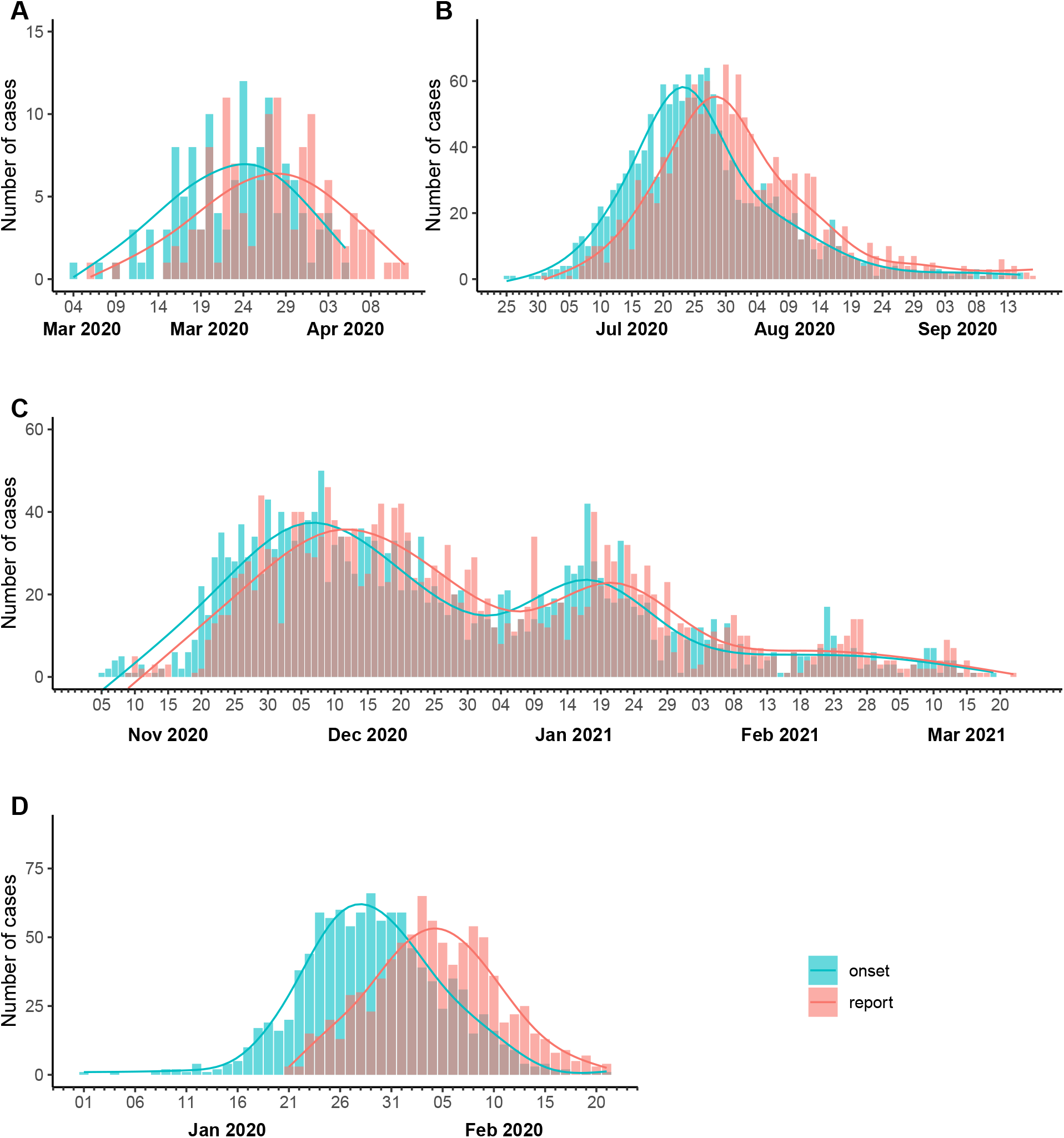
Major epidemic waves in Hong Kong between 1 March 2020 and 23 March 2021 and in mainland China between 1 January to 29 February 2020. The panels represent the (A) second wave, (B) third wave, and (C) fourth wave in Hong Kong, and (D) the first wave in mainland China. The epidemic curves were constructed based on the date of symptom onset (green bars) and the date of the reporting confirmed by RT-PCR test (red bars). The respective solid lines in each panel represent smoothed trends fitted with a generalized additive model.

We constructed case transmission pairs following previously reported methods ^12,30,34^ and estimated *SI* and *SI*_*d*_ distributions (details in the Material and Method section). The distributions of *SI* and *SI*_*d*_ were comparable across epidemic waves, with shorter means for *SI*_*d*_ compared to *SI* (Table S1). In particular, the mean *SI* and *SI*_*d*_ were estimated to be 4.46 (95% CrI: 3.84, 5.08) and 2.59 (95% CrI: 1.28, 3.89) days for the second wave, 3.44 (3.20, 3.68) and 1.93 (1.76, 2.11) days for the third wave, and 3.61 (3.45, 3.76) and 2.18 (2.04, 2.32) days for the fourth wave, respectively. The mean *SI* and *SI*_*d*_ were found to be smaller for the third and fourth waves compared to the initial second wave. Significant variations were observed in the mean *SI* and *SI*_*d*_ both within and across epidemic waves. Similar to the mean *SI*, the mean *SI*_*d*_ was significantly shortened across the pre-, during, and post-peak periods, with shorter mean in post-peak periods for each wave except the second wave (Table S1). The mean estimates of *SI*_*d*_ decreased from 2.80 days (1.32, 4.28) during the pre-peak to 2.72 days (1.34, 4.11) during the post-peak for the second wave, 2.35 days (2.03, 2.67) to 1.40 days (1.11, 1.70) or the third wave; and from 2.81 days (2.51, 3.1) to 1.76 days (1.53, 1.98) for the fourth wave.

We also found that the mean *SI*_*d*_ was significantly shorter than *SI* across most factors and sub-factors for each wave in Hong Kong, except for imported cases (sample size, *n* = 16), short isolation delay *(n* = 1278), tertiary generation infections *(n* = 65), and cases detected in private healthcare facilities *(n* = 598) (Table S1). Moreover, we identified age (≥65 years), disease severity, and delays in isolation/reporting as the significant drivers for both *SI* and *SI*_*d*_ (Figure S2). Notably, isolation and reporting delays exerted opposite effects: longer isolation delays (longer than the median delay) for infectors lengthened *SI* by 0.75-fold while shortened *SI*_*d*_ by 0.35-fold only. The prolonged reporting delays (greater than the median) could shorten *SI*_*d*_ and lengthen *SI*. Local transmission in infectors returned longer *SI*_*d*_ (3.16-fold compared imported cases), while household infections have negative impact on *SI*_*d*_ it by shortening 0.35-fold. Severe cases elevated both metrics *(ST*. +0.48-fold; *S:* +0.48-fold). Similarly, detection methods found to be significant in predicting *SI*_*d*_ with positive impact for private testing but negative for contact tracing. We observed a significant variation in the estimates of *SI* and *SI*_*d*_ across the pre-, during and post-peak each wave.

### Comparing the time-varying inference of SI and SI_d_

Time-varying effective *SI, SI*_*d*_, distributions were estimated using a 10-day sliding window for each wave with their respective uncertainty (95% CrI) (Material and Method section). The distributions were largely comparable, showing similar patterns of change over time. However, daily variations in the estimates of mean effective *SI* and effective *SI*_*d*_ were observed for each of the waves in Hong Kong and mainland China (Figure 2). In Hong Kong, along with the mean effective *SI*, effective *SI*_*d*_ showed daily variations in their estimates, either shortening or lengthening in the same direction (most of the second wave, some phases of the third and fourth waves) or in opposite directions (in certain phases of the third and fourth waves). The mean daily absolute differences between mean effective *SI* and effective *SI*_*d*_ were 0.40 days (IQR: 0.21, 1.03) for the second wave, 1.66 days (IQR: 0.99, 2.31) for the third wave, and 1.42 days (IQR: 0.84, 1.95) for the fourth wave in Hong Kong (Figure 2A-C), and 0.63 days (IQR: 0.29, 1.46) for the first wave in mainland China (Figure 2D).

**Figure 2.**
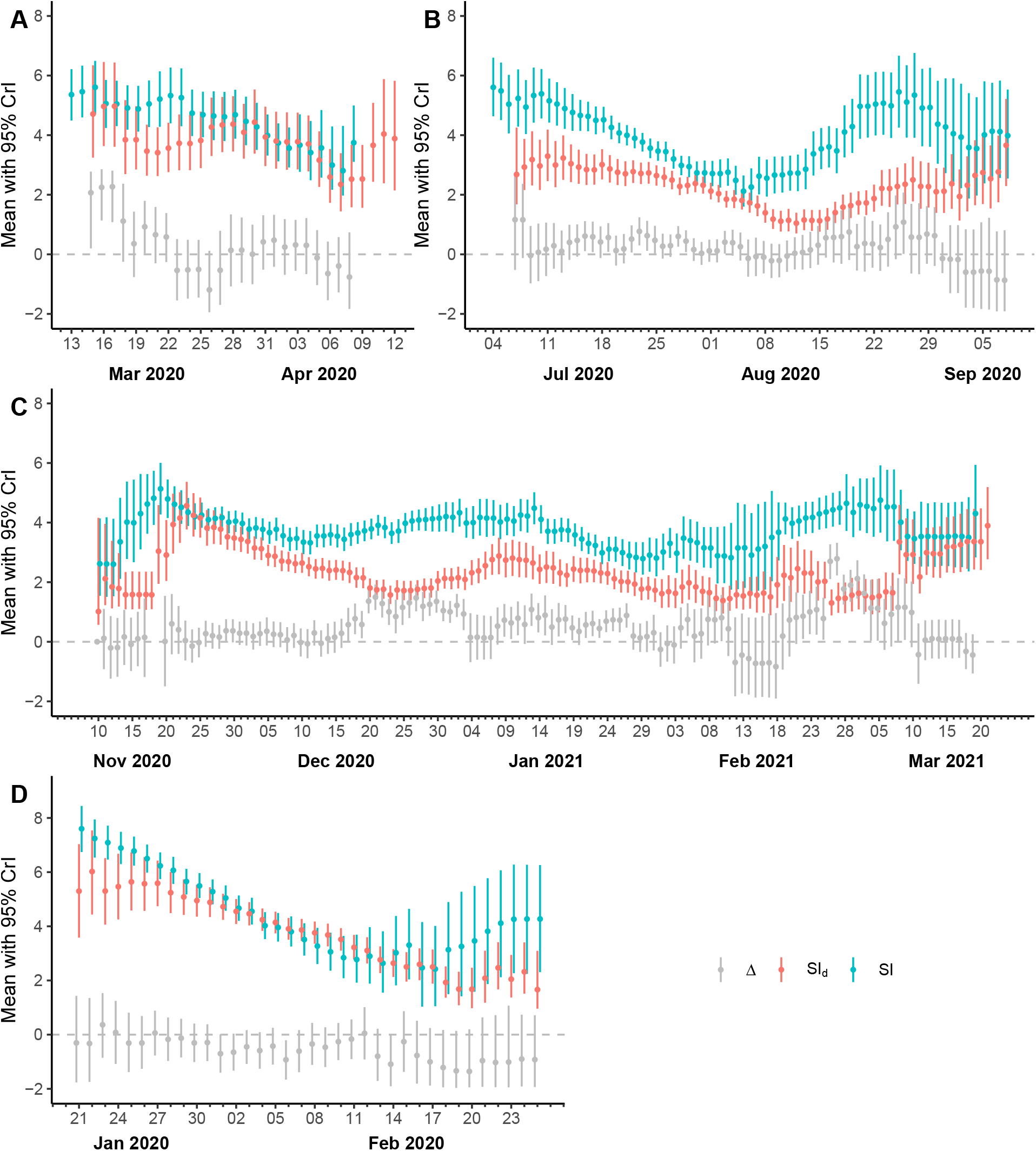
Temporal pattern of diagnostic *(SI*_*d*_) and clinical *(SI)* serial intervals and the differences (Δ) between *d*_1_ (time since onset to report for infectors) and d_2_ (time since onset to report for infectees) for the second (Δ), third (B) and fourth (C) pandemic waves in Hong Kong and first wave in mainland China (D). *SI* was estimated as the interval between successive onsets in the transmission chain and *SI*_*d*_ was the time between diagnostic reporting between cases in transmission pairs. Δ was estimated as the distributions of observed differences between *d*_1_ (onset-to-report delay for infectors) and *d*_*2*_ (onset-to-report delay for infectees). The points in respective colors represent the estimate of means in each time point considering 10-days moving windows and the related error bars are the 95% credible intervals (CrI).

We also evaluated the distributions of observed differences (Δ) between *d*_1_ (onset-to-report delay for infectors) and *d*_*2*_ (onset-to-report delay for infectees) to assess the impact of reporting on *SI*_*d*_ at temporal scale (Figure S1). The distributions were comparable across the waves and locations, with mean differences 0.52 days (IQR: 0.32, 0.76) for the second wave, 0.40 days (0.16, 0.59) for the third wave, and 0.51 days (0.19, 0.85) for the fourth wave in Hong Kong, and 0.46 days (0.30, 0.92) for the first wave in mainland China (**Figure S3**). In general, the temporal estimates of the mean reporting delays *d*_1_ and *d*_*2*_ were found to vary with a longer mean reporting delay for infectors *(df)* compared to infectees *(d*_*2*_) across the waves in Hong Kong, except during the post-peak period of the first wave in mainland China (Figure S3).

Using linear regression model, we further investigated the degree to which differences in *SI*_*d*_ can be attributed to *d*_1_ and *d*_*2*_ (Supplementary text, Section 5). In the base model with only *d*_*1*_ and *d*_*2*_ could explain up to 40% of the variation in *SI*_*d*_ for the second wave (Table S3), while 21% and 23% variation were explained for the third and fourth waves, respectively (Table S4 & S5). After adjustments for other factors, the models could explain up to 92 % of the *SI*_*d*_ variance in the second wave, 69% in the third wave, and 75% in the fourth wave in Hong Kong (Table S3-S5). However, for the combined data of all Hong Kong waves, the explained variation reached a maximum of 53% after adjustments (Table S2).

### SI and SI_d_ on inferring time-varying transmissibility

Using time-varying parametric distributions (means and standard deviations) for both *SI* and *SI*_*d*_, we estimated the time-varying estimates of transmissibility *R*_*t*_ (Material and Method section). We found that the overall trends in *R*_*t*_, derived using both effective *SI* and *SI*_*d*_ distribution for reported epi-curve and onset epi-curve, were comparable with some variation during initial and later phases of the waves in Hong Kong and mainland China (Figure 3). In particular, the inference of mean *R*_*t*_ using proposed effective *SI*_*d*_ distributions and reported epi-curves exhibited closer agreement with *R*, estimates derived from the effective *SI* and onset epi-curve with median absolute difference (MAD) ranged between 0.12 (IQR: 0.08, 0.15) and 0.19 (IQR: 0.10, 0.28) across the location and waves (Table S6). Similarly, estimates of *R*_*t*_ inferred using the effective *SI* and reported epi-curve was comparatively closer to the *R*_*t*_ inferred by *SI*_*d*_ distributions and reported epi-curves with MAD values ranging 0.08 (IQR: 0.04, 0.16) to 0.16 (IQR: 0.12, 0.18). This finding was found to be consistent with other measures, dynamic time warp (DTW) and Euclidean distance (ED) across the waves. Similar results were found when *R*_*t*_ was evaluated under very traditional constant retrospective parametric distributions for *SI* and *SI*_*d*_, and case data from respective epi-curves (Figure S4).

**Figure 3.**
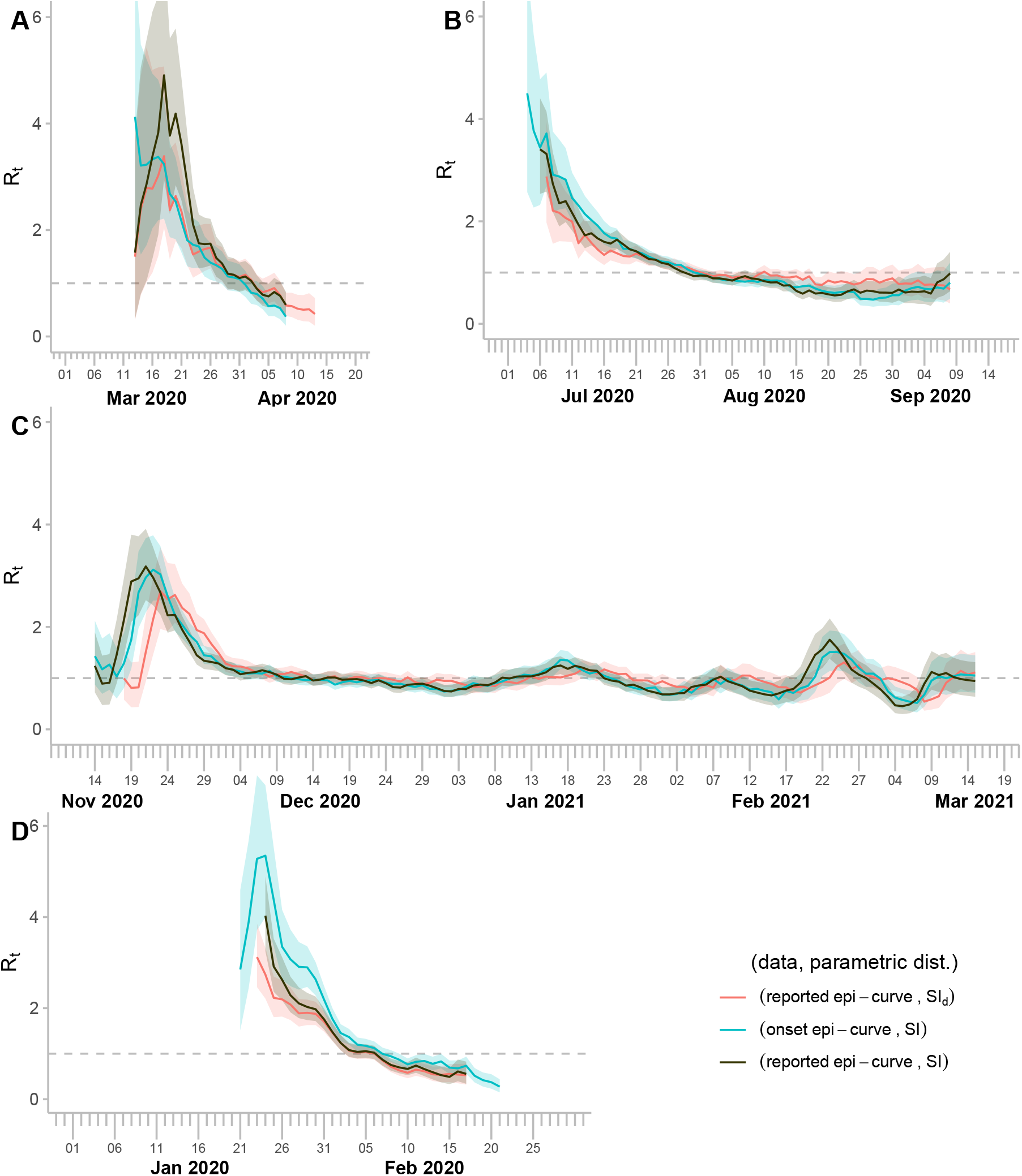
Transmissibility, represented by effective reproduction number *R*_*t*_, of COVID-19 in Hong Kong in the second (A), third (B) and fourth (C) pandemic waves in Hong Kong and first wave in mainland China (D). The red line represents the estimates of *R*_*t*_ based on reported epi-curve and the time-varying diagnostic serial interval *(SI*_*d*_), and the green line was obtained using the onset epi-curve and the time-varying clinical serial interval *(SI)*. The black line shows the traditional method based estimates of *R*_*t*_ using reported epicurve and time-varying *SI*. The respective shaded regions are for 95% confidence intervals. The gray dashed line in each panel represents the threshold value of *R*_*t*_ (= 1) when the outbreak under control in the population.

The sensitivity analysis on earlier reporting (1-4 days earlier) revealed that the time-varying distributions of *SI*_*d*_ in terms of trend and magnitude, were consistent with only minor variation in the mean *SI*_*d*_ and corresponding uncertainty (95% CrI) (Figure S5A). The absolute changes in the *SI*_*d*_ estimates (for 1-4 days earlier over original report dates) ranged between 0% and 19% (Figure S5B), which could lead very minor initial changes in daily *R*_*t*_ estimates (Figure S5C). These findings suggest that the *SI*_*d*_ estimate is not potentially sensitive to changes in the estimation process.

We developed a simulation framework to evaluate the comparability of different epi-curves and parametric distributions in estimating *R*_*t*_ (Material and Method section). Utilizing a susceptible-infectious-recovered (SIR) model, we generated epidemic events and their timings, including case infections, symptom onsets, and reporting dates. This approach enabled us to reconstruct the corresponding epi-curves and the transmission chains between infectors and infectees. The simulation study demonstrated that the time-varying inference of the mean *R*_*t*_ derived from the effective *SI*_*d*_ distribution (using reported epi-curve) were comparable to those obtained via the traditional method (effective *SI* with reported epicurves) under intervention (contract tracing) scenarios with 60% effectiveness (Figure 4A-C). The optimal mean squared differences (MSD) for *R*_*t*_ estimates were calculated relative to reference values derived from infection epi-curve and *GT*. The shortest time to achieve optimal MSD was 6 days for (onset epi-curve, *SI)* combinations and 7 days for (reported epicurve, *SI)* combinations. In contrast, the (reported epi-curve, *SI*_*d*_) combination required 13 days to reach optimal MSD (Figure 4D). Sensitivity analysis confirmed that the timing of optimal MSD remained stable across all parameters-epi-curve combination when tested on 50-100% of datasets (Figure 4E). Results were further validated under additional intervention effectiveness scenarios with 0% (no intervention), 30%, and 90% (Figures S6-S8).

**Figure 4.**
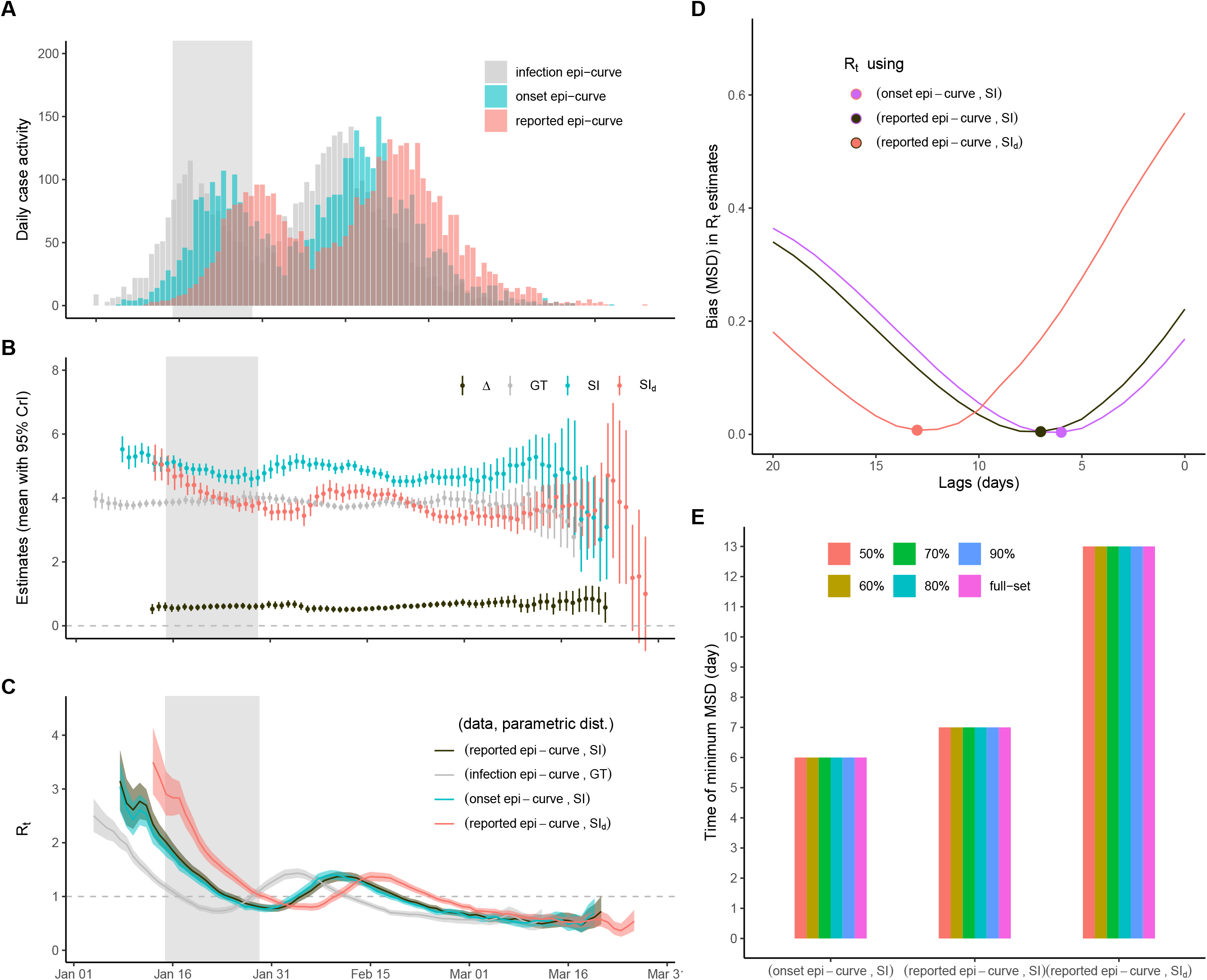
Simulation framework for evaluating the performance of diagnostic serial interval *(SI*_*d*_) in inferring time-varying transmissibility *(R*_*t*_)) by comparing respective biases with respect to the intrinsic transmissibility under specific intervention: early isolation of the cases after onset. (A) Reconstruction of epidemic curves based on the timing of infections, symptom onsets, and reports of cases under 60% intervention effectiveness for early isolation of the cases after onset (additional scenarios in Supplementary Figure. S6-S8). (B) Time-varying estimates of generation time *(GT)*, serial interval *(SI)*, and diagnostic serial interval *(SI*_*d*_) distributions with mean in respective color points and 95% credible intervals (CrI) in the vertical error lines, considering 10-days moving windows. (C) Time-varying reproduction number *(R*_*t*_) estimated by pairing epidemic curves (based on infection, onset, reporting timings) with parametric distributions of infectiousness *(GT, SI, SI*_*d*_). The red line represents the estimates of *R*_*t*_ based on reported epi-curve and the time-varying *SI*_*d*_, and the green line was obtained using the onset epi-curve and the timevarying *SI*. The black line shows the traditional method of estimating *Rt* based on reported epi-curve and time-varying *SI*. The shaded regions were given for 95% confidence intervals. The gray dashed line represented the threshold value of *R*_*t*_ when transmissibility was stable in the population. (D) Biases in respective *R*_*t*_ estimates quantified as mean squared differences (MSD), calculated relative to the reference intrinsic *R*_*t*_ derived from the infection curve and *GT*. Lines indicate the respective biases in *R*_*t*_ estimates for different lags (0-20 days) and dots are the optimal lag time (days) as minimizing MSD. (E) Optimal lag time (days) as minimizing MSD in respective estimated *R*_*t*_ with various length of its timeseries (50% to 100%) to see the sensitivity of the data used. The light-gray shaded region is the timing of the intervention period.

## Discussion

Using contact tracing data on COVID-19 collected between 1 March 2020 and 23 March 2021, encompassing three epidemic waves in Hong Kong and very first wave in mainland China, we inferred the diagnostic serial interval and assessed its performance as an alternative measure to the clinical serial interval. The time-varying inference of *SI* and *SI*_*d*_ could produce the comparable time-varying estimates of *R*_*t*_ evaluated using the reported epi-curve and the onset epi-curve across the waves (Figure 3). The *SI*_*d*_ estimates were found to be mostly shorter compared to the estimates of *SI* across the waves, consistent with the impact of effective contact tracing to identify secondary cases during the initial waves in Hong Kong and mainland China^41^.

In theory, even for given *SI* the estimate of *SI*_*d*_ can vary when the reporting delay for infectors and infectees vary over time (Figure S1). We identified reporting delay as a key factor for the variation in *SI*_*d*_ across the waves (Figure S2). In particular, the variations in reporting delays for both infectors and infectees during the third wave in Hong Kong and the first wave in mainland China reveals a bell-shaped (unimodal) pattern, with the longest delays coinciding with the epidemic peaks (Figure S3). This observed pattern in reporting delay indicates that the surge in the number of cases during these peak periods likely overwhelmed the healthcare and surveillance systems, resulting in significant delays in the reporting process^42^ and can alert the system^41^. Even for given surveillance systems, which could ensure consistent reporting delay for infector and infectee, *SI*_*d*_ can vary based on several factors including PHSMs, case profile and demography and their temporal variations as reported for *SI* in earlier studies ^12,30,31,36^. Along with reporting delays, inclusion of these factors (isolation, severity, household infection, local cases, and contact tracing) in the regression models could explain up to 69%-92% of the variation in *SI*_*d*_ across the waves in Hong Kong, emphasizing their crucial role in shaping transmission dynamics (Table S3-S5). These findings are consistent with mechanistic evidence indicating that early isolation of SARS-CoV-2-infected individuals reduces both generation and serial intervals while increasing the proportion of presymptomatic transmission ^12^’^30^’ ^31^’ ^43^. Notably, the delay between symptom onset and isolation emerged as a dominant determinant of outbreak controllability, particularly at low reproductive numbers *(R*_0_ ≈ 1.5) ^44^. This supports previous modeling studies suggesting that effective containment strategies necessitate high selfisolation rates and efficient contact tracing to reduce the effective reproduction number below 1 in the absence of other measures ^45^. The interplay between imported and local cases significantly shapes transmissibility dynamics, particularly in regions with strong global connections. Initially, in Hong Kong, approximately 70% of total cases were either imported or linked to imported cases ^46^. However, the implementation of stringent control measures, including tightened border controls, quarantine protocols, and enhanced surveillance, played a crucial role in managing the transmissibility of imported cases. These measures effectively reduced the transmissibility of imported cases, maintaining it well below 1 in subsequent waves in Hong Kong ^47^

In scenarios where case numbers are low but the proportion of asymptomatic cases is high as estimated up to 30%-75% for COVID-19 under various settings ^48–51^ potentially lead to sample size limitation, which can introduce biases, particularly early stage of outbreak. In such case, *SI*_*d*_ may be particularly useful, as it accounts for all cases regardless of symptom onset within a given transmission chain. Our analysis of both *SI* and *SI*_*d*_ distributions revealed comparable estimates of *R*_*t*_, with smaller median absolute differences (MAD) values ranging from 0.05 (IRQ: 0.04-0.07) to 0.23 (IQR: 0.13-0.38) across different waves in Hong Kong and mainland China (Figure 3 and Table S6). This disagreement mainly appeared during initial phase of each wave, affected by the different delays in observing onsets and reporting. While reporting delays demonstrated to have impact on *R*_*t*_ estimates, and contribute to the biases in prediction and forecasting of COVID-19 dynamics ^52, 53^. Further temporal variations in reporting delay of confirmed cases significantly affect reported epicurves and hence the estimation of time-varying transmissibility ^54,55^. For earlier relaxation of social distancing policy, *R*_*t*_ raised from 0.7 to 3.2. However, restoration of social distancing measures to its previous level only could reduce *R*_*t*_ to 1.3 due to increased delays in confirmation caused by the additional burden on the contact-tracing system^53^. Our findings indicated that reducing reporting delay especially during active contact tracing significantly shortened effective *SI*_*d*_ (Figure 4B). The degree of shortening of *SI*_*d*_ found to be maximum when intervention effectiveness was at least 60% and beyond, which could substantially impact the effective *R*_*t*_ regardless of derived using *SI* or *SI*_*d*_.

Recent studies illustrated that for estimating *R*_*t*_, forward effective *SI* has been reported as a better parametric distribution for approximating *GT* compared to constant *SI*^12,30,31,36,37^. Our proposed alternative metric, effective *SI*_*d*_ could provide comparable estimates of *R*_*t*_ when using reported epi-curve. Theoretically, *SI*_*d*_ with the reported epi-curve and *SI* with the onset epi-curve can yield reasonable real-time estimates for *R*_*t*_. However, the common practice is to use *SI* with the reported epi-curve, which is ideal for retrospective *R*_*t*_ calculations using constant *SI*. While *SI* is often dependent on self-reporting, using *SI*_*d*_ could serve as a better alternative for real-time *R*_*t*_ estimates when employing the reported epi-curve. Our simulation analysis indicated that time-varying estimates of *R*_*t*_ using intrinsic *GT*, effective *SI* and effective *SI*_*d*_ on the respective epi-curves were comparable with overall trends across the waves, exhibiting only temporal biases when other case profile factors kept fixed (Figures 4 and S6). Variations in case profile factors and case-based interventions may also lead to the magnitudinal biases in *R*_*t*_ specially estimated for real-time even using *SI* ^12,30,31,36,56^.

Estimation of effective reproduction number requires the convolution of case data and the parametric distribution of infectiousness ^8,10,57^, which may vary as epidemic progress ^12,31,37^ Therefore, existing conventional approaches, often rely on simplifying assumptions and approximation for constant infectiousness, should carefully be applied to estimate *R*_*t*_ at realtime ^35,57,58^. For instance, substituting generation times with serial intervals may lead to a biased with a larger variance and underestimates transmissibility for a given growth rate, and overestimats growth rate for a give level of transmissibility neglecting the role of multiple potential infectors, or failing to adjust for censoring effects (particularly during exponential phases) can distort estimates of the generation time distribution, case fatality rates, and ultimately affecting growth rates ^12,24,59,60^. Further, convolution of data (reported epi-curve) and parametric distribution *(SI)* from different levels/timescales (former is based on reporting time and latter one is based on onset timing of the cases) eventually lead to biases for realtime inference of transmissibility, *R*_*t*_. In this study, assimilating data (reported epi-curve) and parametric distribution *(SI*_*d*_) at the same timescale of reporting the cases, we demonstrated how *SI*_*d*_ can offer comparative estimate of *R*_*t*_ over those derived from conventional methods (Figure S7 & S8). In general, having *R*_*t*_ based on reported epi-curve and effective *SI*_*d*_, *R*_*t*_ can be further adjusted for temporal biases using time varying incubation period and reporting delay from onset to reconstruct the intrinsic transmissibility at real-time. This finding suggests that *R*_*t*_ based on *SI*_*d*_*-* may offer a practical alternative in settings where generation time and onset time data are scarce or unreliable, provided uncertainties from observation delays and data quality are explicitly modeled.

Our study demonstrated that the diagnostic serial interval could serve as a potential alternative to the clinical serial interval, offering comparable time-varying estimates of transmissibility. Also, the performance of the proposed matric *SI*_*d*_ in *R*_*t*_ estimation validated by the details COVID-19 pandemic data in Hong Kong and mainland China, generalization of this method can be possible for other infectious diseases and across locations, allowing improved surveillance and case detection practices at community level. However, several limitations should be acknowledged. First, to infer *SI*_*d*_, the contact tracing data for transmission chain recontraction is required, which is often not usual to observe for seasonal respiratory viruses including influenza and even for post-pandemic SARS-CoV-2. Second, *SI*_*d*_ is influenced by the efficiency and speed of the detection and reporting system, which can vary depending on the testing strategies employed (e.g., rapid antigen tests (RATs) versus PCR). For instant, during the early stages of the pandemic in Hong Kong, RATs were not widely available^25^; instead, suspected cases were primarily confirmed through PCR testing of nasal or throat samples collected by healthcare professionals^26^. Consequently, delays in detection and reporting can introduce bias in to *SI*_*d*_ estimates. This limitation is particularly pertinent in contexts where healthcare systems are overwhelmed or testing capacity is limited. Third, the proposed measure and its performance were validated using simulation by comparing reconstructed intrinsic transmissibility and the transmissibility under various data and parametric distributions, which can be assessed by real observed data for inferring the time-varying *GT*, predicted from exposure/infection time using contact tracing and viral load data ^12,61^. Finally, while our findings support the use of *SI*_*d*_ as an alternative to *SI*, further research is needed to validate these results in diverse settings and to investigate the potential impact of factors such as vaccination status, variant transmissibility, and changes in public health measures on *SI*_*d*_.

Despite these limitations, our study highlights the potential utility of *SI*_*d*_ as an alternative to *SI* for estimating the effective reproduction number and other transmissibility measures for COVID-19 and other respiratory viruses for given healthcare system. By incorporating both symptomatic and asymptomatic cases, *SI*_*d*_ may offer a more comprehensive understanding of disease transmission characteristics while addressing the limitations and biases associated with self-reported symptom onset data. Further research is essential to validate and refine the application of *SI*_*d*_ across various settings and populations, as well as to explore its potential role in monitoring and controlling the spread of COVID-19 and other infectious diseases.

## Materials and Methods

### Data source and defining onsets of the COVID-19 waves

We retrieved records of all locally diagnosed COVID-19 cases reported in Hong Kong during the first 4 waves until 23 March 2021, from the Department of Health of the Hong Kong SAR government. The records included information such as age, sex, date of symptom onset for symptomatic cases, date of case reporting, and date of isolation or hospital admission. Additionally, cases were categorized as locally infected or imported, based on whether infection occurred within a household or elsewhere (from contact tracing data). Clinical severity outcomes (critical, severe or stable), and the place of testing for detection were also documented. Data from the first COVID-19 wave in Hong Kong, which occurred between January and February 2020, were excluded as most cases were imported with sporadic and limited local transmissions. Cases from all subsequent waves until 23 March 2021, were included, with their respective defined onsets as the “second” wave (1 March 2020 to 10 April 2020), “third” wave (25 June 2020 to 8 September 2020), and “fourth” wave (1 November 2020 to 23 March 2021) ^30^. By analyzing epi-curves for illness onset and reporting dates of the cases, we identified the peak timings for these three waves: from 16 to 24 March 2020 for the second wave, from 18 to 27 July 2020 for the third wave, and the fourth wave had two peaks, the first from 30 November 2020 to 15 December 2020, and the second during 11 to 25 January 2021 (Supplementary text, Section 1) ^30,31^. The fifth wave of COVID-19 in Hong Kong, which began in early January 2022^32^, posed a challenge to contact tracing capacity due to a rapid increase in cases. Therefore, the fifth and all subsequent waves are excluded from the analysis. Similar line-list (contact tracing) data was also retrieved for mainland China, reported by China’s municipal health commissions outside Hubei province from 1 January to 29 February 2020. This data was originally sourced from publicly available case reports provided by over 200 municipal health commissions in mainland China and has been previously reported in other studies (Supplementary text, Section 1)^12, 31, 33, 34^. The linelist data includes the demographic information, exposure and contact history, and potential transmission links.

### Reconstruction of transmission pairs and their factor-based stratifications analyses

Using the relevant line-list (contact tracing) information on confirmed COVID-19 cases, we constructed case transmission pairs following previously reported methods ^12,30,34^ We explored three main criteria: i) recent travel history (to exclude overseas infection), ii) spatial and temporal associations with large local clusters, and iii) assessing whether the workplace was in an area with a high likelihood of contact. In cases of large clusters with complex epidemiological links, cases were individually traced to determine the most probable chains of transmission. For clusters with two or more likely infectors, the case with the earliest onset date was considered the most probable infector in a pair. If paired cases shared the same onset date, the case with the earliest report date or case number would be classified as the infector, and such pairs were termed as probable/likely pairs^12,30,31,35^.

Considering various factors reported to address the variation in the estimate of clinical *SI* ^30,31,36,37^, we explored the impact of these factors on *S* by stratifying the transmission pairs data at temporal and non-temporal settings (rationale and stratification details are in section 1 and 2 in Supplementary text). The stratifications included classifying the transmission pairs for inter-wave, pre-, during, and post-peak within wave, as well as generation status (primary, secondary, and tertiary etc.) as the temporal factors. Other factors were assessed including case demographics such as age (< 65 and ≥ 65 years) and sex, case severity profile^38^ (severe and non-severe), household and non-household infection, source of infections (local and imported), detection setting (private, public, and tracing), case isolation delay (shorter/ longer compare to median isolation delay) of infectors, and reporting delay (shorter/ longer compare to median reporting delay) of infectors. Based on the available information for the mainland, we could consider factors such as age, sex, and household versus non-household infection.

### Estimation of SI_d_, SI, and onset-to-reporting interval distributions

We used Bayesian inferential methods to estimate the mean and standard deviation (sd) with 95% credible intervals (CrI) for *SI, SI*_*d*_, and onset-to-report delay by fitting a series of distributions (e.g., normal and gamma) to the respective empirical data based on observed pairs. Markov chain Monte Carlo (MCMC) methods were employed to estimate posterior distributions, with likelihood functions derived from these distributions. The mean and standard deviation were predicted by taking the median of the means and standard deviation, respectively. The 95% Crl bounds were defined by the 2.5% and 97.5% quantiles (see Supplementary text, Section 3).

To compare impact of different factors, we evaluated these estimates for each factor-based stratified data (as described in above subsection). Independent-sample t-tests were conducted to determine the statistical significance of differences in means between *SI* and *SI*_*d*_ within each stratified category. Additionally, we employed a series of linear regression models to investigate the association of these factors on *SI* and *SI*_*d*_ (Supplementary text, Section 4). We then explored the temporal variations in the estimates of *SI, SI*_*d*_, and onset-to-report by evaluating them across the waves (for Hong Kong only) and pre-, during and post-peak for each wave in Hong Kong and mainland China. Time-varying effective *SI, SI*_*d*_, distributions were estimated using a 10-day sliding window (7-and 14-day windows for sensitivity analysis), for each wave with their respective uncertainty (95% CrI). We also evaluated the distributions of observed differences (Δ) between *d*_*1*_ (onset-to-report delay for infectors) and *d*_*2*_ (onset-to-report delay for infectees) to assess the impact of reporting on *SI*_*d*_ at temporal scale (Figure S1).

Further, we conducted a counterfactual analysis to examine the sensitivity of the timing of epidemic events on *SI*_*d*_ estimates, and how the earlier reporting of an infection would affect the *SI*_*d*_ estimates. To illustrate this, we considered the second wave in Hong Kong to simulate the scenarios where reporting dates for both infectors and infectees were imputed randomly by shifting 1−4 days earlier. Using these imputed dates, we estimated the mean *SI*_*d*_ and 95% CrI for each counterfactual scenario. The percentage change in mean *SI*_*d*_ relative to the original reporting dates was calculated to quantify the impact of reporting timing.

### Estimating effective reproduction number (R_t_) by using SI and SI_d_

We estimated the time-varying effective reproduction number (*R*_*t*_)a measure of transmissibility defined as the average number of secondary infections generated by a single infected individual at time t. Using the estimated time-varying effective *SI* and *SI*_*d*_ distributions, we estimated *R*_*t*_ via the *EpiEstim* package in R^10,39^, which incorporates the daily incidence of infections *(I*_*t*_) weighted by the average of infectiousness, characterized by generation time distributions, approximated by *SI* or *SI*_*d*_ here (Supplementary text, Section 6). Using time-varying parametric distributions (means and standard deviations) for both *SI* and *SI*_*d*_, we estimated *R*_*t*_ estimates.

In practice, the true infection epi-curve and generation time distribution are often unobserved, the reported number of cases (reported epi-curve) and the *SI* distribution were used to estimate *R*_*t*_. However, *SI* distributions rely on symptom onset dates, whereas reported incidence curves depend on reporting dates. To assess how these data and parametric approximations could affect the estimates of transmissibility ***(R***_***t***_), we analyzed the detailed data in Hong Kong and mainland China. We compared the series of ***R***_***t***_ estimates, derived using different available epi-curves and parametric distributions: reported epi-curve paired with effective ***SI*** distributions, onset-based epi-curves paired with effective ***SI*** distributions and reported epi-curve paired with effective ***SI***_***d***_.

### Individual based simulation for transmission chain reconstruction to assess the biases in estimating effective reproduction number (R_t_) by using SI and SI_d_

We developed a simulation framework to assess the comparability of these different epicurves and parametric distributions in estimating *R*_*t*_. We used a susceptible-infectious-recovered (SIR) model to generate the epidemic events and their timing, including case infections, onsets, and reports, which further allowed us to reconstruct respective epi-curves and the infector-infectee transmission chain. We assumed an intrinsic generation time follows a gamma distribution with mean 3.95 days and standard deviation 2.28 days ^40^ and the incubation period *(IP)* follows a gamma distribution with a mean of 5.2 days and standard deviation 1.80days ^40^. Additionally, we considered a fixed mean reporting delay of 6 days, based on the observed mean reporting delay in Hong Kong. Using the transmission pairs data, we first estimated the time-varying distributions of *GT, SI*, and *SI*_*d*_, which allowed us to evaluate the corresponding time-varying *R*_*t*_, based on infection, onset, and reported epicurves respectively.

We further checked the effectiveness of public health and social measures (PHSMs) to reduce the transmissibility using *GT, SI*, and *SI*_*d*_. To incorporate intervention measures, we allowed the epidemic to progress for two weeks during its exponential phase. Following this period, we implemented interventions with contact tracing for early detection of cases with varying levels of effectiveness: 0% (no intervention), 30%, 60%, and 90% reduction in onset-to-confirmation delay. Such intervention could mitigate the outbreaks by restricting forward transmissions in community and allowed to last for two weeks. Using the above simulation framework, we evaluated *R*_*t*_ for intervention scenarios under different combinations of epicurves and parameter distributions: (reported epi-curve, *SI)*, (reported epi-curve, *SI*_*d*_), (onset epi-curve, *SI)* and (infection epi-curve and *GT”)*. The bias in these *R*_*t*_ estimates was assessed by calculating the mean squared differences (MSD) by comparing the *R*_*t*_ under these combinations with the *R*_*t*_ derived from the infection epi-curve and intrinsic *GT*. Further, to validate the framework in detecting temporal biases in *R*_*t*_, we allowed 0 to 20 days lag in the time series of *R*_*t*_ and calculate the MSD, which could be minimum around the assumed intrinsic delay *(IP* = 5.2 days and onset-to-report = 6 days). We also considered different epidemic-onset thresholds (using 50%, 60%, 70%, 80%, 90%, and the full set of data across the peak of each wave) on estimating MSD to identify the sensitivity of variation in *R*_*t*_ during low cases (initial and end phases). All statistical analyses were conducted using R version 4.2.1.

## Data Availability

Statistical analyses were conducted using R version 4.0.5 (R Foundation for Statistical Computing, Vienna, Austria). All data generated from publicly available sources, codes and materials will be provided upon specific request to the author.

## Acknowledgements

The authors thank Julie Au for technical assistance. This project was supported by the University of Hong Kong Grant (project code 2201100868); Health and Medical Research Fund (project no. 20190712); AIR@InnoHK administered by Innovation and Technology Commission, and by a grant from the Research Grants Council of the Hong Kong Special Administrative Region, China (Project No. T11-705/21-N).The funding bodies had no role in study design, data collection and analysis, preparation of the manuscript, or the decision to publish.

## Contributors

All authors meet the International Committee of Medical Journal Editors criteria for authorship. STA and BJC conceived the study, designed the statistical and modelling methods; MPH, AY, JX, DC and STA did the data collection, assimilation and performed the data analysis; MPH, and STA wrote the first draft of the manuscript. All authors contributed to the interpretation of the results, revision of the manuscript critically for intellectual content and have given final approval of the version to be published.

## Supplementary Material

Supplementary information

