## Supplementary file for "Diagnostic serial interval as an alternative measure of clinical serial interval using ancestral COVID-19 waves in Hong Kong and mainland China"

### 1. Defining epidemic waves and temporal stratifications

Our analysis of COVID-19 epidemic waves in Hong Kong focused on the second, third, and fourth waves, excluding the initial wave (23 January–29 February 2020) due to limited local case data and its comparatively milder impact. The second wave (1 March–10 April 2020) exhibited a peak between 16–24 March 2020, during which 53% of cases (34/64) showed symptom onset. The third wave (25 June–8 September 2020) peaked from 18–27 July 2020, accounting for 37% of cases (274/736) in that period. The prolonged fourth wave (1 November 2020–23 March 2021) featured two distinct peaks: 30 November–15 December 2020 and 11–25 January 2021. Collectively, these peaks represented 46% of cases (524/1131) with symptom onset. Peaks were defined as intervals during which ≥25% of confirmed cases exhibited symptom onset, with pre- and post-peak phases determined accordingly ^1^.

In mainland China, our study of the first wave (1 January–18 February 2020) included 428 infectors (individuals transmitting the virus) with symptom onset and 629 transmission pairs. The symptom onset peak for infectors occurred during 23–29 January 2020, encompassing over one-third of cases. A subsequent peak for infectees (individuals contracting the virus) followed approximately one week later (29 January–3 February 2020), with 231 symptomatic infectees (over one-third of the total). Epidemic phases were categorized as pre-peak (1–23 January), peak-timing (23–29 January), and post-peak (29 January–29 February 2020). Analyses were restricted to the first wave due to the unavailability of contact tracing data for subsequent waves; further methodological details on transmission pairs are described in prior work ^2-5^.

### 2. Other factor-based stratifications for the diagnostic serial intervals

Case pairs were categorized by their generational position within transmission chains: primary (directly linked to the index case), secondary (infected by primary cases), and tertiary (infected by secondary cases), with subsequent generations classified analogously. To characterize transmission dynamics, pairs were further stratified by contact setting into household and non-household infections. Household transmission encompassed individuals cohabiting in shared residences, including family members and roommates. Non-household transmission involved exposures in settings such as housing estates (non-cohabiting residents), restaurants, schools, workplaces, and social gatherings.

Cases were additionally classified by detection method into four groups: (1) community (identified via the Universal Community Testing Program [UCTP] or community testing centers); (2) public (detected through general outpatient clinics [GOPC] or enhanced public laboratory surveillance); (3) private (diagnosed in private clinics, via private testing, or through private-sector surveillance systems); and (4) tracing (meeting reporting criteria during medical surveillance). Imported cases were distinguished from local transmissions, with the latter including close contacts of imported, local, or potentially local cases. Finally, age was analyzed as a binary variable (below 65 years vs. 65 years or older) to assess demographic risk patterns.

### 3. Defining the effective diagnostic serial interval, effective clinical serial interval, onset-to-reporting interval distributions and their inference

We quantified three key epidemiological intervals: (1) the diagnostic serial interval, defined ($SI_{d}$) as the time between reports in transmission chain (Supplementary Figure. S1); (2) reporting delay, the time between the symptom onset to reporting of the case; and (3) isolation delay, the time from symptom onset to isolation of the case. Reporting and isolation delays were further categorized as "short" or "long" relative to their respective median values. Specifically, delays shorter than the median were classified as short, while those exceeding the median were designated as long.

To estimate $SI_{d}$, we employed Bayesian methods, modeling the time difference between reporting dates of infectors ($T_{r1}$) and infectees ($T_{r2}$) as $SI_{d}= T_{r2} - T_{r1}$. The model incorporated the number of transmission pairs and reporting dates, assuming a normal likelihood function parameterized by the mean ($\mu$) and standard deviation ($\sigma$) of $SI_{d}$ (Equation 1):

$$p\left( y \right|\mu, \sigma) = \left( \frac{1}{\sigma\sqrt{\left( 2 \pi\right)}} \right) exp\left( -\frac{\left( y -\mu\right)^{2}}{2 \sigma^{2}} \right)$$

where $y$ represents the observed $SI_{d}$ values. Priors for $\mu$ and $\sigma$ were specified as a normal distribution (Equation 2) and gamma distribution (Equation 3), respectively:

$$p(\mu) = \left( \frac{1}{\sigma_{\mu} \sqrt{2 \pi}} \right)exp\left( -\frac{\left( \mu-\mu_{0} \right)^{2}}{2 \sigma_{\mu}^{2}} \right)$$

and

$$p(\sigma) = \left( \frac{b^{a}}{\Gamma\left( a \right)} \right)\sigma^{a - 1}exp(-b\sigma)$$

with prior mean ($\mu_{0}$), prior standard deviation ($\sigma_{\mu}$), gamma shape ($a$) and rate ($b$) parameters, and gamma function ($\Gamma\left( a \right)$). Posterior distributions were computed via Bayes’ theorem (Equation 4):

$$p(\mu, \sigma| y) = p(y \left| \mu, \sigma\right)\times p(\mu) p(\sigma)$$

Posterior sampling was performed using Markov chain Monte Carlo (MCMC) methods implemented in the ***rstan*** package. Analogous frameworks were applied to estimate clinical serial intervals ($SI = T_{o2} - T_{o1}$, where $T_{o}$ denotes symptom onset dates) and onset-to-confirmation delays for infectors ($d_{1} = T_{r1} - T_{o1}$) and infectees ($d_{2} = T_{r2} - T_{o2}$).

For each epidemic wave, we calculated effective diagnostic serial intervals ($SI_{d}$), clinical serial intervals ($SI$), and onset-to-confirmation delays $(d_{1}, d_{2}$). Temporal trends were analyzed using 10-day moving windows, with means and 95% credible intervals (CrI) estimated at each time point. Differences between $SI$ and $SI_{d}$ estimates were visualized to assess temporal shifts in transmission dynamics and potential statistical significance.

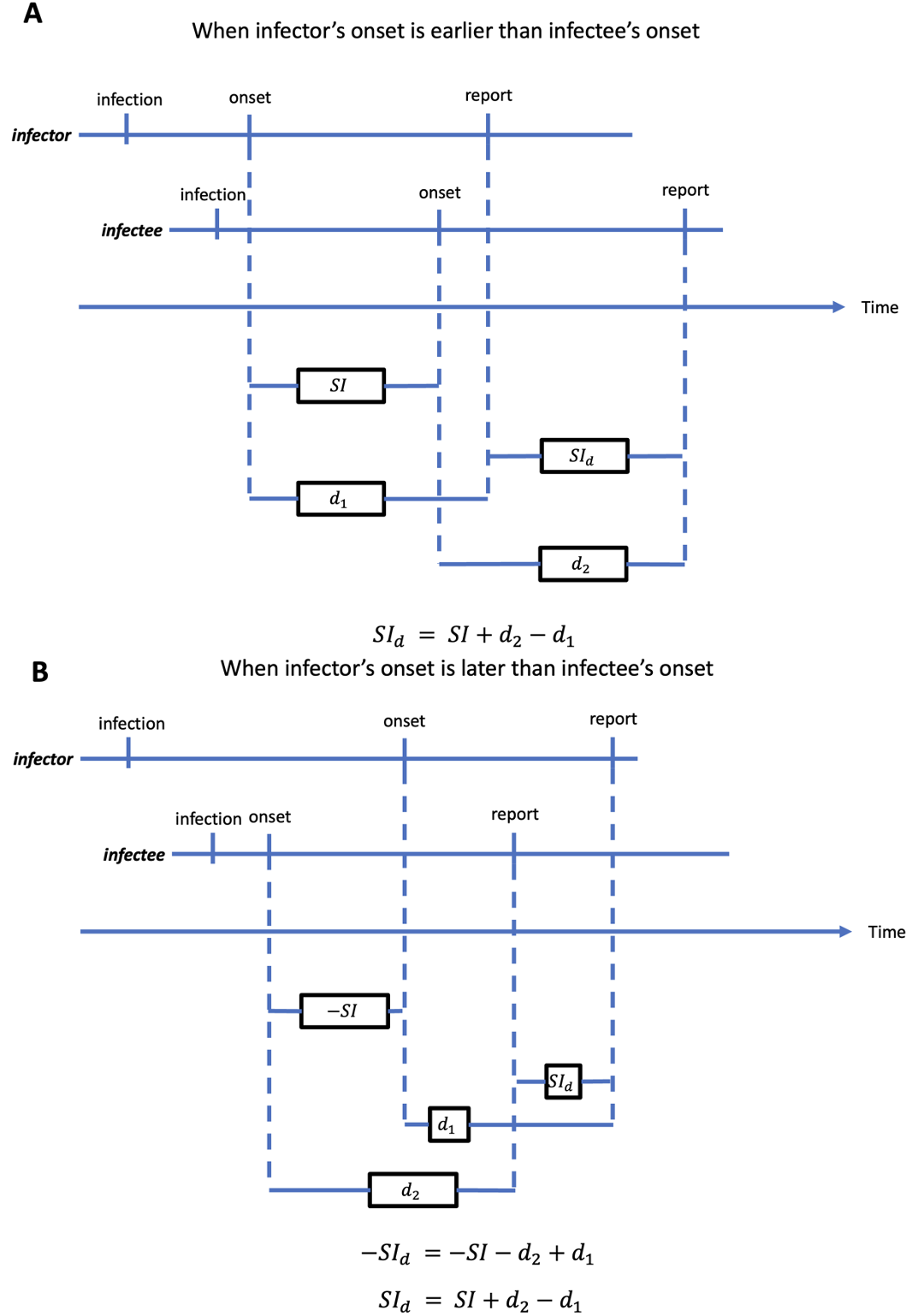

**Figure S1**. Schematic illustration of the relationship between $SI$ and $SI_{d}$. (A) Relationship between $SI$ and $SI_{d}$ when infector’s date of onset is earlier than the infectee’s date of onset. (B) Relationship between $SI$ and $SI_{d}$ when infector’s date of onset is later than the infectee’s date of onset.

**Table S1**. **Stratified mean clinical and diagnostic serial interval estimates are presented with their corresponding 95% confidence intervals**. The mean interval is assumed to follow a normal distribution. The p-value was calculated using a two-sample t-test. The null hypothesis tested was that there is no difference between the mean $SI$ and $SI_{d}$ for each stratified category.

| **Variables** | | **Second wave** | | | **Third wave** | | | **Fourth wave** | | | **Overall** | | |
| --- | --- | --- | --- | --- | --- | --- | --- | --- | --- | --- | --- | --- | --- |
|  |  | **Mean** $\boldsymbol{SI}$ **(95% CrI)** | **Mean** $\mathbf{S}\mathbf{I}_{\mathbf{d}}$ **(95% CrI)** | **p-value** | **Mean** $\boldsymbol{SI}$ **(95% CrI)** | **Mean** $\mathbf{S}\mathbf{I}_{\mathbf{d}}$ **(95% CrI)** | **p-value** | **Mean** $\boldsymbol{SI}$**(95% CrI)** | **Mean** $\mathbf{S}\mathbf{I}_{\mathbf{d}}$ **(95% CrI)** | **p-value** | **Mean** $\boldsymbol{SI}$ **(95% CrI)** | **Mean** $\mathbf{S}\mathbf{I}_{\mathbf{d}}$ **(95% CrI)** | **p-value** |
| **Overall** | | 4.46(3.84-5.08) | 2.59(1.28-3.89) | 0.0111 | 3.44(3.2-3.68) | 1.93(1.76-2.11) | 0.0000 | 3.61(3.45-3.76) | 2.18(2.04-2.32) | 0.0000 |  |  |  |
| **Age** | |  |  |  |  |  |  |  |  |  |  |  |  |
|  | <= 65 | 4.46(3.84-5.08) | 2.59(1.28-3.89) | 0.0111 | 3.29(3.02-3.57) | 1.8(1.61-2) | 0.0000 | 3.49(3.32-3.66) | 2.15(2-2.3) | 0.0000 | 3.45(3.31-3.6) | 2.03(1.91-2.16) | 0.0000 |
|  | > 65 | - | - | - | 4.04(3.52-4.56) | 2.45(2.03-2.87) | 0.0000 | 4.11(3.76-4.47) | 2.3(1.94-2.65) | 0.0000 | 4.08(3.78-4.38) | 2.36(2.09-2.63) | 0.0000 |
| **Gender** | |  |  |  |  |  |  |  |  |  |  |  |  |
|  | female | 4.26(3.27-5.26) | 3.71(2.35-5.06) | 0.5002 | 3.61(3.26-3.96) | 1.94(1.68-2.19) | 0.0000 | 3.6(3.36-3.84) | 2.3(2.08-2.51) | 0.0000 | 3.62(3.42-3.82) | 2.19(2.03-2.36) | 0.0000 |
|  | male | 4.58(3.77-5.4) | 1.87(-0.1-3.84) | 0.0128 | 3.27(2.93-3.6) | 1.93(1.68-2.17) | 0.0000 | 3.62(3.42-3.82) | 2.07(1.89-2.25) | 0.0000 | 3.52(3.35-3.7) | 2(1.84-2.17) | 0.0000 |
| **Source** | |  |  |  |  |  |  |  |  |  |  |  |  |
|  | imported | 4.17(2.18-6.15) | 1.83(-10.44-6.78) | 0.1605 | - | - | - | 1.25(0.45-2.05) | 2.25(-1.73-6.23) | 0.4861 | 3.44(1.85-5.03) | -0.81(-7.1-5.48) | 0.1806 |
|  | local | 4.51(3.84-5.17) | 3.29(2.48-4.11) | 0.0232 | 3.44(3.2-3.68) | 1.93(1.76-2.11) | 0.0000 | 3.61(3.46-3.77) | 2.18(2.04-2.32) | 0.0000 | 3.57(3.44-3.71) | 2.11(2.01-2.22) | 0.0000 |
| **Setting** | |  |  |  |  |  |  |  |  |  |  |  |  |
|  | household | 4.76(3.82-5.7) | 3.74(2.64-4.84) | 0.1564 | 3.55(3.28-3.83) | 1.84(1.65-2.03) | 0.0000 | 3.59(3.41-3.76) | 2.03(1.88-2.18) | 0.0000 | 3.6(3.45-3.75) | 1.99(1.87-2.11) | 0.0000 |
|  | non-household | 4.18(3.34-5.02) | 1.51(-0.8-3.83) | 0.0333 | 3.05(2.54-3.56) | 2.24(1.81-2.67) | 0.0171 | 3.67(3.34-4.01) | 2.68(2.34-3.02) | 0.0001 | 3.48(3.2-3.75) | 2.43(2.12-2.73) | 0.0000 |
| **Severity** | |  |  |  |  |  |  |  |  |  |  |  |  |
|  | non-severe | 4.23(3.63-4.83) | 2.48(1.05-3.91) | 0.0265 | 3.29(3.02-3.55) | 1.87(1.67-2.07) | 0.0000 | 3.56(3.39-3.73) | 2.19(2.04-2.34) | 0.0000 | 3.48(3.34-3.62) | 2.08(1.95-2.21) | 0.0000 |
|  | severe | 6.2(3.19-9.21) | 3.4(0.02-6.78) | 0.1787 | 4.19(3.62-4.75) | 2.23(1.86-2.61) | 0.0000 | 3.93(3.5-4.36) | 2.09(1.71-2.46) | 0.0000 | 4.12(3.77-4.47) | 2.19(1.92-2.46) | 0.0000 |
| **Detection** | |  |  |  |  |  |  |  |  |  |  |  |  |
|  | private | 2.33(0.9-3.77) | 2(-0.48-4.48) | 0.6495 | 4.18(3.73-4.63) | 2.59(2.3-2.89) | 0.0000 | 3.7(3.42-3.98) | 2.65(2.37-2.93) | 0.0000 | 3.9(3.65-4.15) | 2.62(2.42-2.82) | 0.0000 |
|  | public | 4.87(3.46-6.28) | 2.26(1.06-3.46) | 0.0055 | 3.18(2.89-3.46) | 1.89(1.66-2.11) | 0.0000 | 3.75(3.47-4.02) | 2.09(1.88-2.3) | 0.0000 | 3.46(3.26-3.66) | 1.98(1.83-2.14) | 0.0000 |
|  | Tracing | 4.41(3.69-5.13) | 2.74(0.91-4.56) | 0.0923 | 2.48(1.4-3.57) | 0.1(-0.56-0.76) | 0.0003 | 3.54(3.09-3.98) | 1.33(0.9-1.76) | 0.0000 | 3.44(3.05-3.83) | 1.29(0.85-1.73) | 0.0000 |
| **Isolation delay** | |  |  |  |  |  |  |  |  |  |  |  |  |
|  | long-delay | 5.57(4.48-6.66) | 0.83(-2.11-3.77) | 0.0037 | 4.4(4.06-4.73) | 1.64(1.41-1.87) | 0.0000 | 4.47(4.21-4.73) | 1.05(0.85-1.25) | 0.0000 | 4.47(4.26-4.69) | 1.36(1.18-1.55) | 0.0000 |
|  | short-delay | 3.71(3.03-4.4) | 3.77(2.85-4.69) | 0.9196 | 2.28(1.95-2.6) | 2.28(2.01-2.55) | 0.9915 | 3.16(2.96-3.37) | 2.84(2.65-3.04) | 0.0234 | 2.88(2.71-3.05) | 2.69(2.53-2.84) | 0.0958 |
| **Reporting delay** | |  |  |  |  |  |  |  |  |  |  |  |  |
|  | long-delay | 5.4(4.41-6.39) | 1.12(-1.48-3.73) | 0.0031 | 4.61(4.25-4.98) | 1.5(1.25-1.75) | 0.0000 | 4.46(4.22-4.69) | 0.89(0.71-1.07) | 0.0000 | 4.57(4.36-4.78) | 1.18(1-1.36) | 0.0000 |
|  | short-delay | 3.66(2.93-4.39) | 3.83(2.88-4.78) | 0.7763 | 2.42(2.12-2.72) | 2.31(2.07-2.55) | 0.5552 | 3.15(2.95-3.34) | 2.88(2.7-3.05) | 0.0431 | 2.91(2.74-3.07) | 2.71(2.56-2.85) | 0.0667 |
| **Generation** | |  |  |  |  |  |  |  |  |  |  |  |  |
|  | primary | 4.58(3.91-5.25) | 2.4(0.96-3.83) | 0.0071 | 3.49(3.22-3.75) | 1.85(1.66-2.04) | 0.0000 | 3.66(3.49-3.82) | 2.06(1.91-2.21) | 0.0000 | 3.62(3.48-3.77) | 1.99(1.86-2.11) | 0.0000 |
|  | secondary | 3.75(2.09-5.41) | 4.62(1.91-7.34) | 0.5282 | 3.27(2.55-4) | 2.6(2.17-3.04) | 0.1199 | 3.35(2.84-3.86) | 2.84(2.46-3.21) | 0.1104 | 3.33(2.92-3.74) | 2.8(2.52-3.09) | 0.0371 |
|  | tertiary + | - | - | - | 2.26(0.59-3.93) | 2.16(0.87-3.45) | 0.9171 | 3.07(2.23-3.9) | 3.27(2.46-4.07) | 0.7290 | 2.8(2.06-3.54) | 2.91(2.24-3.57) | 0.8292 |
| **Peak timing** | |  |  |  |  |  |  |  |  |  |  |  |  |
|  | pre-peak | 5.50(4.41-6.59) | 2.80(1.32-4.28) | 0.0040 | 4.57(4.13-5.02) | 2.35(2.03-2.67) | 0.0000 | 4.07(3.78-4.35) | 2.81(2.51-3.1) | 0.0000 | 4.32(4.07-4.56) | 2.62(2.4-2.83) | 0.0000 |
|  | during peak | 4.51(3.61-5.41) | 2.45(0.21-4.68) | 0.0903 | 3.14(2.81-3.47) | 2.01(1.72-2.31) | 0.0000 | 3.30(3.07-3.54) | 2.03(1.83-2.24) | 0.0000 | 3.30(3.11-3.49) | 2.05(1.86-2.24) | 0.0000 |
|  | post-peak | 3.17(1.99-4.34) | 2.72(1.34-4.11) | 0.6086 | 2.67(2.19-3.16) | 1.40(1.11-1.7) | 0.0000 | 3.60(3.31-3.89) | 1.76(1.53-1.98) | 0.0000 | 3.20(2.94-3.46) | 1.63(1.45-1.81) | 0.0000 |

### 4. Potential factors for the variation in $\boldsymbol{SI}$ and $\boldsymbol{S}\boldsymbol{I}_{\boldsymbol{d}}$

### We examined the relationships between driving factors of $\boldsymbol{SI}$ and $\boldsymbol{S}\boldsymbol{I}_{\boldsymbol{d}}$ using a generalized

### linear regression model with an identity link function and Gaussian family. The model was

### formulated as:

$$y=\alpha+\sum_{i=1}^{p} \beta_{i}x_{i}+\epsilon$$

Here, $y$ represents either $SI$ or$SI_{d}$, $\alpha$ denotes the intercept, $\beta_{i}$ corresponds to the coefficient of predictor $x_{i}$, and $x_{i}$ encompasses the following covariates: age categories (<65 vs. ≥65 years), sex (female vs. male), infection source (imported vs. local), transmission setting (non-household vs. household), disease severity (non-severe vs. severe), detection method (community-based, private, public, or contact tracing), isolation delay (short vs. long), reporting delay (short vs. long), case generation (primary vs. secondary), and peak timing (pre-peak vs. peak; pre-peak vs. post-peak).

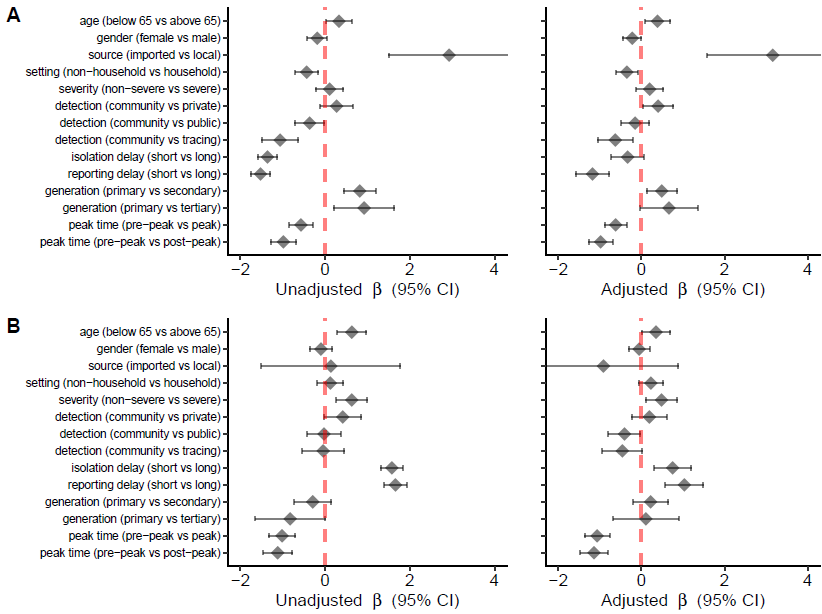

**Figure S2.** Association between (A) diagnostic serial interval ($SI_{d}$) and its driving factors, and (B) clinical serial interval ($SI$) and its driving factors.

### 5. Comparing the temporal distributions of $\boldsymbol{d}_{\boldsymbol{1}}$ and $\boldsymbol{d}_{\boldsymbol{2}}$ and their impact on $\boldsymbol{S}\boldsymbol{I}_{\boldsymbol{d}}$

Marginally distinct distributions were observed for the differences (Δ) between reporting delays for infectors ($d_{1}$) and infectees ($d_{2}$) across various epidemics waves. Generally, the temporal patterns in the mean reporting delays ($d_{1}$ and $d_{2}$) were analogous, with infectors ($d_{1}$) experiencing longer mean reporting delays compared to infectees ($d_{2}$) throughout the waves in Hong Kong, except during the post-peak period of the first wave in mainland China

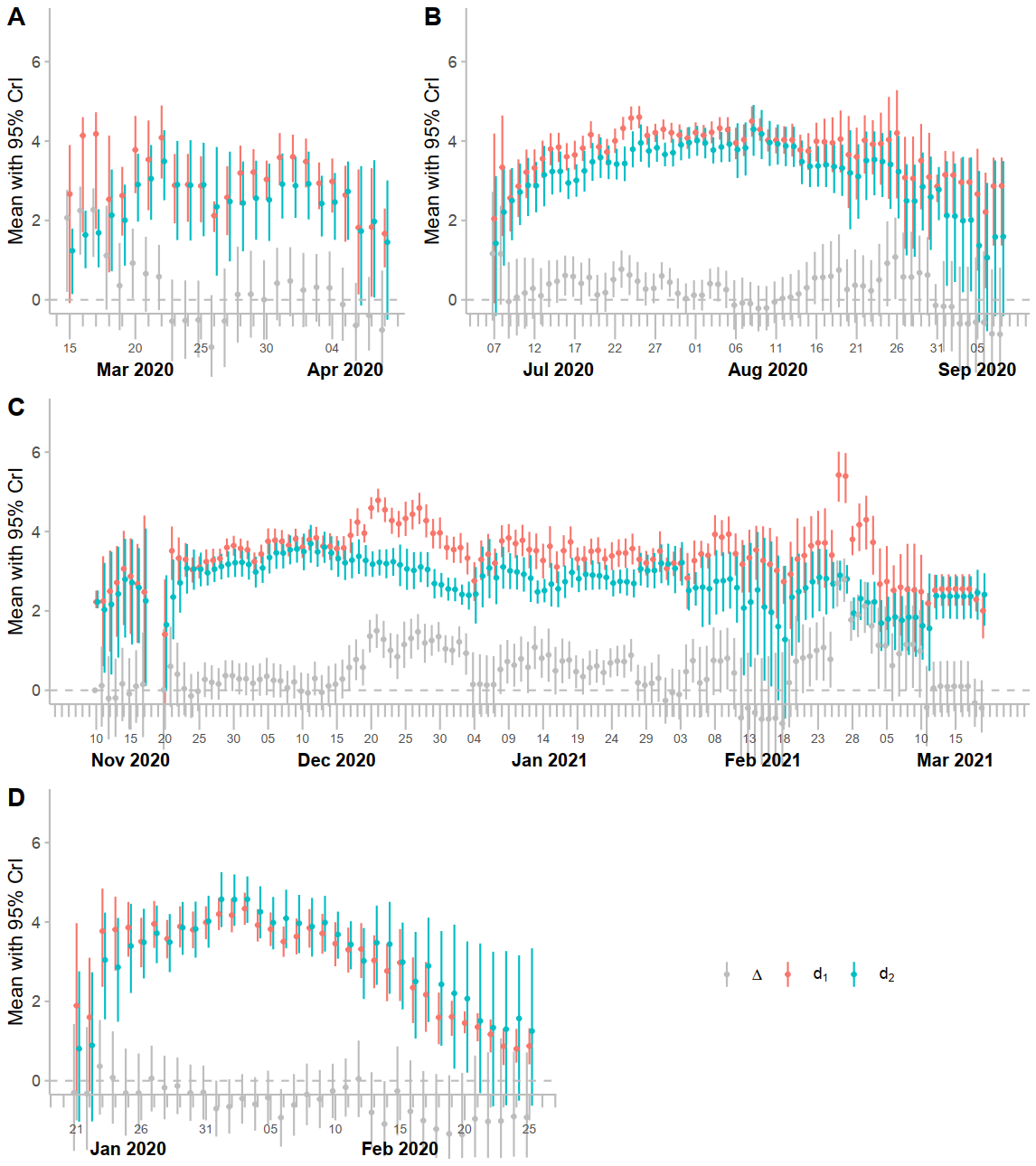

**Figure S3.** Temporal pattern of reporting delay (difference between onset and reporting) among the infectors ($d_{1}$) and infectee ($d_{2}$) and their differences (Δ) in the second, third and fourth epidemic waves in Hong Kong (A-C), respectively and the first wave in Mainland China (D). The points represent the estimate of means in each time point considering 10-days moving windows and the error bars depict the 95% credible intervals (CrI).

Theoretical equality is observed between $SI$ and $SI_{d}$, as well as between $d₁$ and $d₂$ (Figure S1). However, the extent to which differences in $SI_{d}$ are attributable to $d₁$ and $d₂$ requires deeper exploration. To investigate this, we performed a linear regression analysis using data from Hong Kong’s second, third, and fourth epidemic waves. The base model was formulated as follows:

$$SI_{d} = \alpha+ \beta_{1} d_{1} + \beta_{2} d_{2} + \epsilon$$

Subsequent adjustments to this model incorporated additional covariates, including the proportion of local cases ($L$), household transmission ($H$), severe cases ($S$), cases identified via contact tracing ($T$), cases with prolonged isolation delays ($D$), and cases reported during epidemic peaks ($P$). When aggregating data across all three waves, the base model explained only 7% of the total variation in $SI_{d}$ (Table S2). However, after adjusting for the aforementioned factors, the explained variation increased by 53%, highlighting the collective influence of these variables.

Further stratification by epidemic wave revealed distinct patterns. The base model applied to the second wave explained significantly more variation (40%) in $SI_{d}$ compared to the third (21%) and fourth waves (23%) (Table S4). Strikingly, after adjustment for covariates, $d₁$ and $d₂$ accounted for 91% of the variation in $SI_{d}$, underscoring the critical role of these parameters in explaining differences across waves when contextual factors are considered.

**Table S2**. Explained variation of $SI_{d}$ by $d_{1}$ (difference between onset and reporting among the infectors) and $d_{2}$ (difference between onset and reporting among the infectees) **considering data in 2^nd^, 3^rd^ and 4^th^ waves in Hong Kong**. Multiple factors were considered for adjustments, such as, proportion of local cases (L), proportion infection in household settings (H), proportion sever cases (S), proportion of cases identified by contract tracing (T), proportion of cases with long isolation delay (D), proportion of cases reported during the peak time of epidemic (P).

| **Factors** | **Coeff of** $\boldsymbol{d}_{\boldsymbol{1}}$**:** $\boldsymbol{\beta}_{\boldsymbol{1}}$ | **Coeff of** $\boldsymbol{d}_{\boldsymbol{2}}$**:** $\boldsymbol{\beta}_{\boldsymbol{2}}$ | **Adj.** $\boldsymbol{R}^{\boldsymbol{2}}$ | $\boldsymbol{\Delta}\boldsymbol{R}^{\boldsymbol{2}}$ |
| --- | --- | --- | --- | --- |
| d1+d2 | -0.388 | 0.0334 | 0.073 | - |
| d1+d2+T | -0.383 | 0.235 | 0.307 | 0.234 |
| d1+d2+T+D | -0.149 | 0.176 | 0.464 | 0.062 |
| d1+d2+T+D+L | -0.182 | 0.269 | 0.526 | 0.062 |
| **d1+d2+T+D+L+S** | **-0.171** | **0.261** | **0.529** | **0.234** |
| d1+d2+T+D+L+S+H | -0.176 | 0.265 | 0.527 | 0.062 |

**Table S3**. Explained variation of $SI_{d}$ by $d_{1}$ (difference between onset and reporting among the infectors) and $d_{2}$ (difference between onset and reporting among the infectees) **considering data in 2^nd^ wave in Hong Kong**. Multiple factors were considered for adjustments, such as, proportion of local cases (L), proportion infection in household settings (H), proportion sever cases (S), proportion of cases identified by contract tracing (T), proportion of cases with long isolation delay (D), proportion of cases reported during the peak time of epidemic (P).

| **Wave** | **Factors** | **Coeff of** $\boldsymbol{d}_{\boldsymbol{1}}$**:** $\boldsymbol{\beta}_{\mathbf{1}}$ | **Coeff of** $\boldsymbol{d}_{\boldsymbol{2}}$**:** $\boldsymbol{\beta}_{\mathbf{2}}$ | **Adj.** $\boldsymbol{R}^{\mathbf{2}}$ | $\boldsymbol{\Delta}\boldsymbol{R}^{\boldsymbol{2}}$ |
| --- | --- | --- | --- | --- | --- |
| 2nd wave | d1+d2 | 0.6959 | -0.4837 | 0.397 | - |
| 2nd wave | d1+d2+D | -0.0101 | 0.3413 | 0.739 | 0.342 |
| 2nd wave | d1+d2+D+S | 0.2266 | -0.1311 | 0.823 | 0.084 |
| 2nd wave | d1+d2+D+S+H | 0.4373 | -0.4027 | 0.853 | 0.030 |
| 2nd wave | d1+d2+D+S+H+L | 0.4856 | -0.4935 | 0.875 | 0.022 |
| **2nd wave** | **d1+d2+D+S+H+L+T** | **0.2434** | **-0.3502** | **0.918** | **0.043** |
| 2nd wave | d1+d2+D+S+H+L+T+P | 0.2726 | -0.3911 | 0.914 | -0.004 |

**Table S4**. Explained variation of $SI_{d}$ by $d_{1}$ (difference between onset and reporting among the infectors) and $d_{2}$ (difference between onset and reporting among the infectees) **considering data in 3^rd^ wave in Hong Kong**. Multiple factors were considered for adjustments, such as, proportion of local cases (L), proportion infection in household settings (H), proportion sever cases (S), proportion of cases identified by contract tracing (T), proportion of cases with long isolation delay (D), proportion of cases reported during the peak time of epidemic (P).

| **Wave** | **Factors** | **Coeff of** $\boldsymbol{d}_{\boldsymbol{1}}$**:** $\boldsymbol{\beta}_{\mathbf{1}}$ | **Coeff of** $\boldsymbol{d}_{\boldsymbol{2}}$**:** $\boldsymbol{\beta}_{\mathbf{2}}$ | **Adj.** $\boldsymbol{R}^{\mathbf{2}}$ | $\boldsymbol{\Delta}\boldsymbol{R}^{\boldsymbol{2}}$ |
| --- | --- | --- | --- | --- | --- |
| 3rd wave | d1+d2 | 0.2991 | -0.6054 | 0.207 | - |
| 3rd wave | d1+d2+D | 0.2780 | -0.5219 | 0.583 | 0.376 |
| 3rd wave | d1+d2+D+H | 0.4882 | -0.7556 | 0.659 | 0.076 |
| 3rd wave | d1+d2+D+H+P | 0.4957 | -0.8795 | 0.686 | 0.027 |
| 3rd wave | d1+d2+D+H+P+S | 0.4533 | -0.8970 | 0.688 | 0.002 |
| **3rd wave** | **d1+d2+D+H+P+S+L** | **0.4533** | **-0.8970** | **0.688** | **0.000** |
| 3rd wave | d1+d2+D+H+P+S+L+T | 0.3954 | -0.9240 | 0.686 | -0.002 |

**Table S5**. Explained variation of $SI_{d}$ by $d_{1}$ (difference between onset and reporting among the infectors) and $d_{2}$ (difference between onset and reporting among the infectees) **considering data in 4^th^ wave in Hong Kong**. Multiple factors were considered for adjustments, such as, proportion of local cases (L), proportion infection in household settings (H), proportion sever cases (S), proportion of cases identified by contract tracing (T), proportion of cases with long isolation delay (D), proportion of cases reported during the peak time of epidemic (P).

| **Wave** | **Factors** | **Coeff of** $\boldsymbol{d}_{\boldsymbol{1}}$**:** $\boldsymbol{\beta}_{\mathbf{1}}$ | **Coeff of** $\boldsymbol{d}_{\boldsymbol{2}}$**:** $\boldsymbol{\beta}_{\mathbf{2}}$ | **Adj.** $\boldsymbol{R}^{\mathbf{2}}$ | $\boldsymbol{\Delta}\boldsymbol{R}^{\boldsymbol{2}}$ |
| --- | --- | --- | --- | --- | --- |
| 4th wave | d1+d2 | -0.6257 | 0.7728 | 0.229 | - |
| 4th wave | d1+d2+P | -0.6065 | 1.1793 | 0.403 | 0.174 |
| 4th wave | d1+d2+P+L | -0.7733 | 1.2255 | 0.538 | 0.135 |
| 4th wave | d1+d2+P+L+D | -0.3582 | 0.7115 | 0.617 | 0.079 |
| 4th wave | d1+d2+P+L+D+T | -0.2745 | 0.6742 | 0.675 | 0.058 |
| 4th wave | d1+d2+P+L+D+T+H | -0.3435 | 0.8886 | 0.729 | 0.054 |
| **4th wave** | **d1+d2+P+L+D+T+H+S** | **-0.3623** | **0.9086** | **0.753** | **0.024** |

### 6. Estimating effective reproduction number by using $\boldsymbol{SI}$ and $\boldsymbol{S}\boldsymbol{I}_{\boldsymbol{d}}$

The real-time transmissibility of an infectious disease is quantified by the effective reproduction number ($R_{t}$), presents the expected number of secondary infections generated by an infector on a given day ^6,7^. $R_{t}$ is considered under control when it is less than 1, indicating that the spread of the pathogen is being contained, while values greater than 1 suggest ongoing transmission. To estimate $R_{t}$, we utilized a stable serial interval distribution to approximate the generation time distribution. Specifically, we employed $SI$ and $SI_{d}$ as a proxies. The daily estimate of $R_{t}$ is calculated by dividing the number of new cases ($I_{t}$) on a given day by the weighted average of infectiousness caused by cases infected on previous days, as described by the following formula: $R_{t}=\frac{I_{t}}{\sum_{i=1}^{t} I_{t-i}w_{i}}$, where $w_{i}$ represents the serial interval distribution ^8^. $R_{t}$ is equal to $I_{t}$ divided by the sum of $I_{t-i}$ multiplied by $w_{i}\left( t \right)$ for the time-varying clinical serial interval, and by $f_{i}\left( t \right)$ for the $SI_{d}$. The probability that individuals generate secondary infections at time $t$, given that they were confirmed $i$ days before, is described by $f_{i}\left( t \right)$. Conversely, $w_{i}\left( t \right)$ describes the probability of individuals generating secondary infections at time $t$, given that they were infected $i$ days prior.

**Table S6.** The dynamic time warp (DTW), Euclidean distance (ED) and median absolute difference (MAD) of $R_{t}$ time series estimated using effective $SI_{d}$ distribution and reported epi-curves, effective $SI$ and onset epi-curve, and reported epi-curve and effective $SI$.

| **Waves and locations** | **Epi-curves and parameters combination** | **Reported epi-curve,** $\boldsymbol{S}\boldsymbol{I}_{\boldsymbol{d}}$  Median (IQR) | | |
| --- | --- | --- | --- | --- |
|  |  | **MAD** | **ED** | **DTW** |
| **Hong Kong**  **(Wave 2)** | (reported epi-curve, $SI$) | 0.08  (0.04, 0.16) | 3.27  (1.93-4.41) | 4.48 |
|  | (onset epi-curve, $SI$) | 0.19  (0.10, 0.28) | 2.99  (1.08, 4.87) | 5.12 |
| **Hong Kong**  **(Wave 3)** | (reported epi-curve, $SI$) | 0.16  (0.12, 0.18) | 1.55  (1.31, 1.78) | 7.27 |
|  | (onset epi-curve, $SI$) | 0.14  (0.09, 0.18) | 2.30  (1.79, 2.83) | 6.01 |
| **Hong Kong**  **(Wave 4)** | (reported epi-curve, $SI$) | 0.13  (0.09, 0.16) | 3.67  (2.18, 4.99) | 13.42 |
|  | (onset epi-curve, $SI$) | 0.12  (0.08, 0.15) | 4.48  (2.51, 6.20) | 15.89 |
| **Mainland China**  **(Wave 1)** | (reported epi-curve, $SI$) | 0.11  (0.09, 0.12) | 0.82  (0.58, 1.04) | 1.62 |
|  | (onset epi-curve, $SI$) | 0.17  (0.15, 0.22) | 1.02  (0.89, 1.14) | 1.75 |

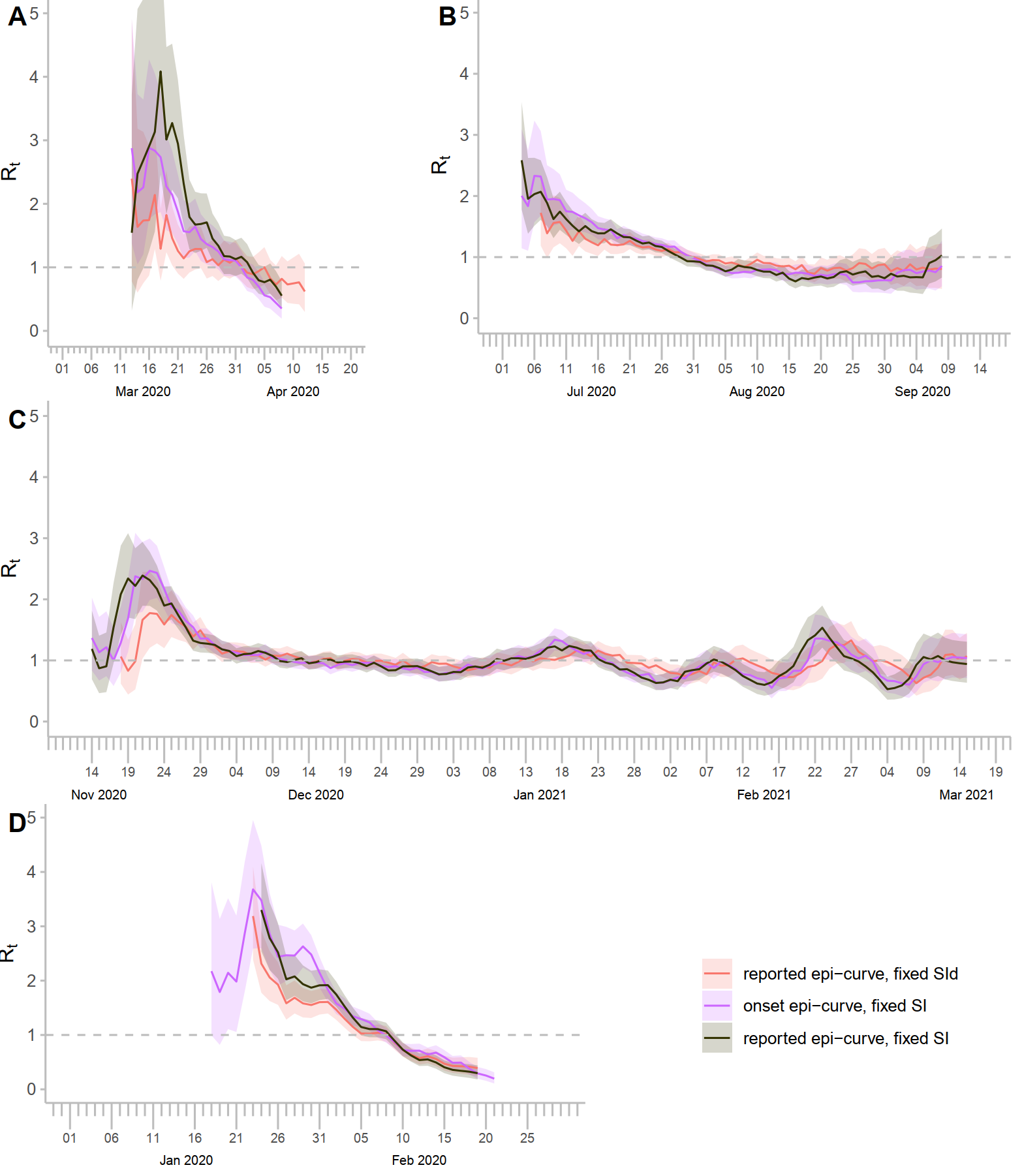

**Figure S4.** Transmissibility, represented by effective reproduction number *R_t_*, of COVID-19 in Hong Kong in the second, third and fourth epidemic waves (A-C), respectively and (D) the first wave in mainland China. The red line represents the estimates of *R_t_* based on reported epi-curve and the mean diagnostic serial interval ($SI_{d}$), and the green line was obtained using the onset epi-curve and the mean clinical serial interval (*SI*). The black line shows the traditional method of estimating *R_t_* based on reported epi-curve and *SI*. The shaded regions were given for 95% confidence intervals. The gray dashed line in each panel represented the threshold value of *R_t_* when transmissibility was stable in the population.

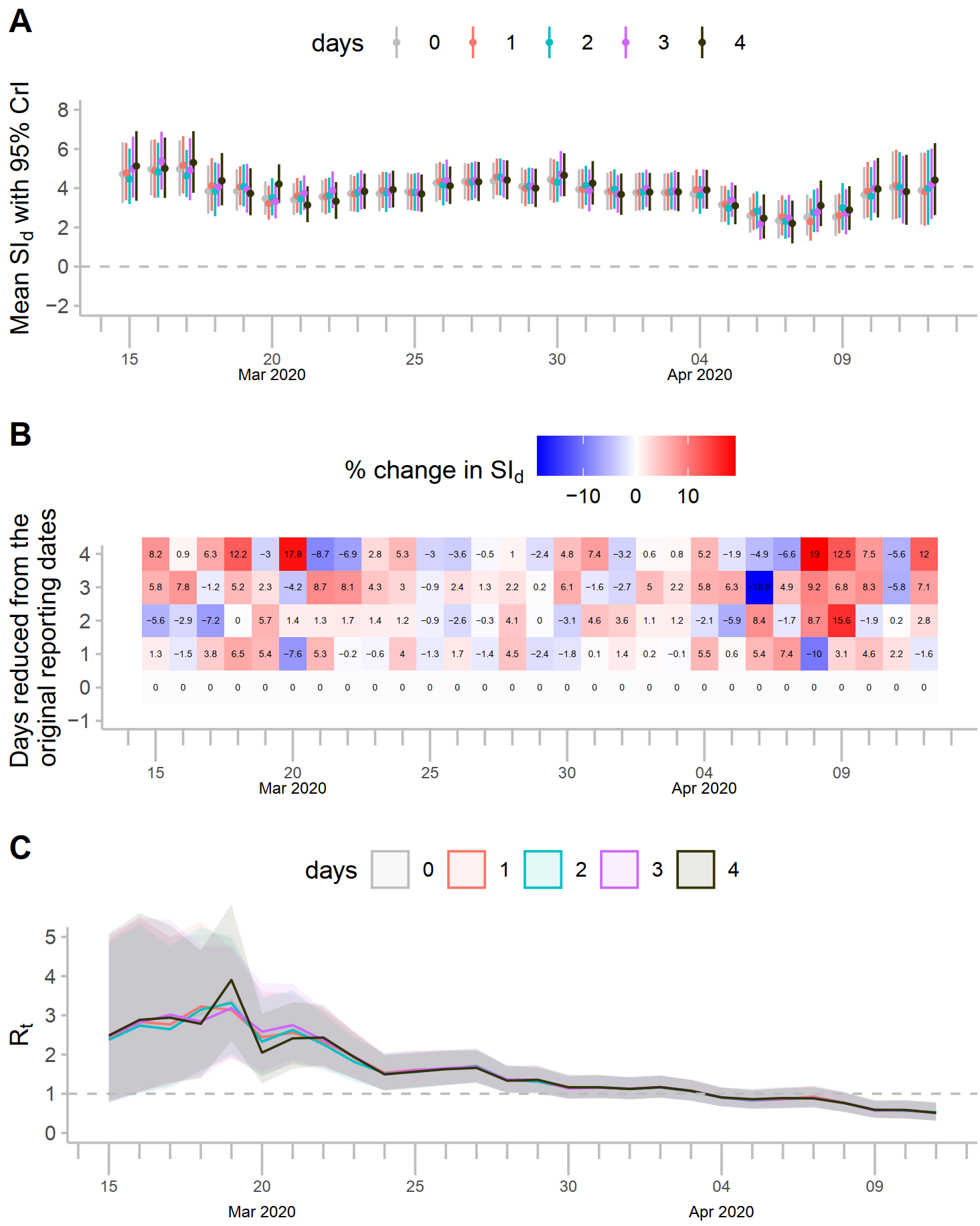

**Figure S5**. Sensitivity analysis of the $SI_{d}$ estimates using data from the second wave in Hong Kong. (A) Time-varying estimates of $SI_{d}$ if the reporting is made earlier, between 0-4 days, where 0 days represent the original reporting date, and 4 indicates if reporting was made four days earlier. Dots represent the mean value of $SI_{d}$, and the bars represent the 95% CrI. (B) Heatmap represents the percentage change in $SI_{d}$ estimate with reference to the original reporting dates. (C) The estimated effective reproduction number for varying reductions in reporting dates.

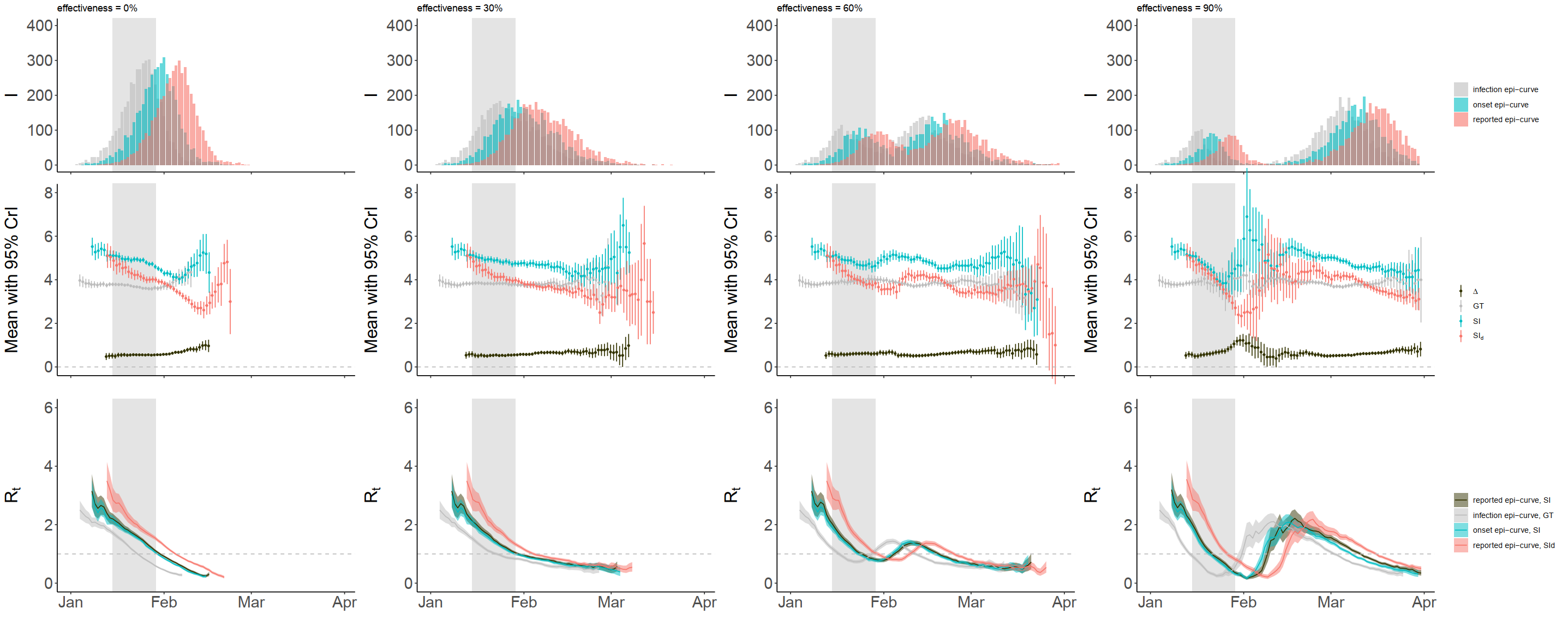

**Figure S6**. The epidemic wave was simulated under the scenario (from left to right) with intervention (contract tracing) effectiveness 0% (no intervention), 30%, 60% and 90% respectively. Interventions were implemented starting on day 14 of the outbreak and maintained for 14 consecutive days in all scenarios. (Top Panel): Simulated epidemic wave. The epidemic curve was simulated using a fixed $R_{0}$ of 2.5 (as estimated elsewhere ^9^) and mean reporting delay of 6 days (observed mean reporting delay in Hong Kong ) from the symptom onset. (Middle Panel): Temporal estimates of $SI_{d}$ (red colour), $SI$ (blue colour), and their differences (grey colour). The dots represent the mean values, while the error bars indicate the 95% credible intervals (CrI). The grey dots with error bars depict the mean differences between $d_{1}$ and $d_{2}$ along with their corresponding 95% CrI. (Bottom Panel): $R_{t}$ of COVID-19. The shaded regions indicate the 95% confidence intervals. The grey dashed line in each panel represents the threshold value of $R_{t}$ when transmissibility is stable in the population.

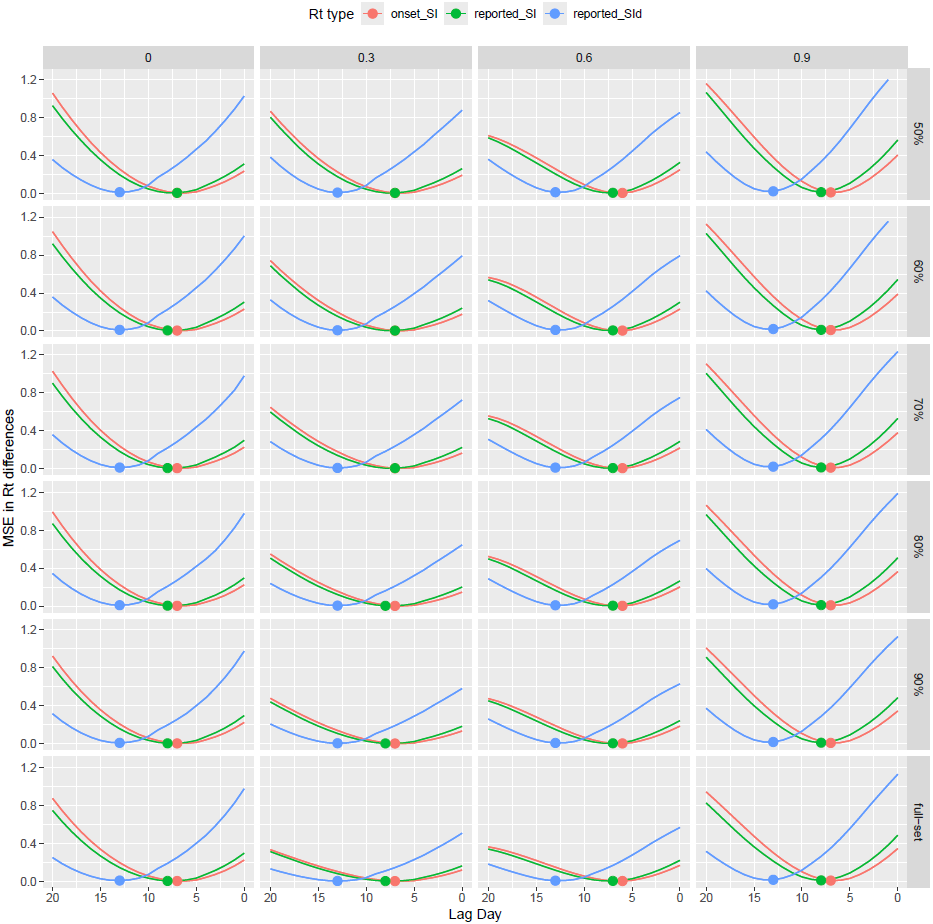

**Figure S7**. Mean squared differences (MSD) in $R_{t}$ differences. Differences were calculated with reference to the $R_{t}$ estimates based on the infection epi-curve and generation time ($GT$). Lines represent the MSD, and points indicate the minimum MSD. The $x$-axis represents the lag days relative to the original $R_{t}$ estimates. Each column of panels presents different scenarios of intervention effectiveness, including 0% (no intervention), 30%, 60%, and 90% effectiveness. Each row of panels indicates the different threshold values considered for estimating the $R_{t}$ differences, specifically 50%, 60%, 70%, 80%, 90%, and the full set of data.

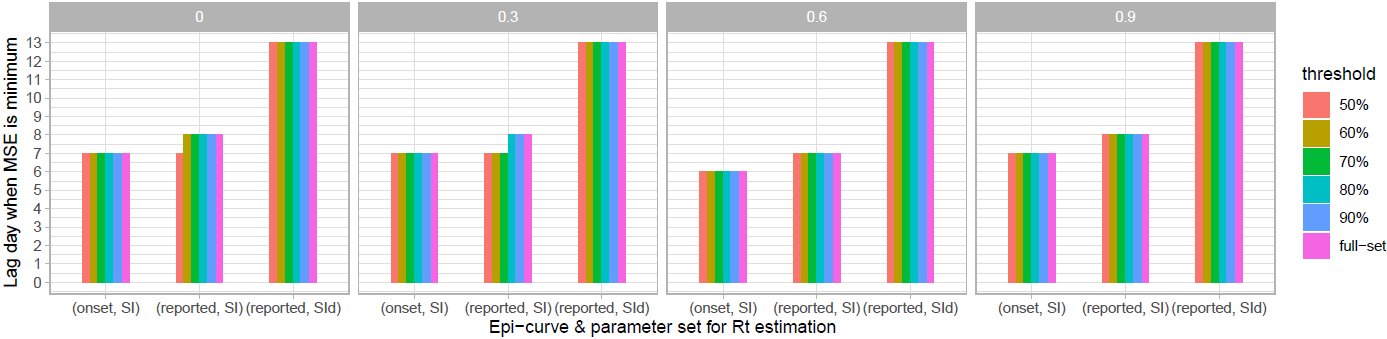

**Figure S8**. Lag days when the mean squared differences (MSD) of $R_{t}$ were at their minimum. The MSD was calculated as the mean of the squared differences between $R_{t}$ estimates based on the infection epi-curve and $GT$, and $R_{t}$ estimates based on other combinations of epi-curves and parameter distributions. Each panel displays estimates for intervention effectiveness at 0% (no intervention), 30%, 60%, and 90% for various threshold values.
